# Histology of type 3 macular neovascularization and microvascular anomalies in treated age-related macular degeneration: a case study

**DOI:** 10.1101/2022.09.13.22279910

**Authors:** Andreas Berlin, Diogo Cabral, Ling Chen, Jeffrey D Messinger, Chandrakumar Balaratnasingam, Randev Mendis, Daniela Ferrara, K. Bailey Freund, Christine A Curcio

## Abstract

**Objective/Purpose:** To investigate intraretinal neovascularization and microvascular anomalies by correlating in vivo multimodal imaging with corresponding ex vivo histology in a single patient.

**Design:** A case study comprising clinical imaging from a community-based practice, and histologic analysis at a university-based research laboratory (clinicopathologic correlation).

**Participants:** A white woman in her 90’s treated with numerous intravitreal anti-vascular endothelial growth factor (VEGF) injections for bilateral type 3 macular neovascularization (MNV) secondary to age-related macular degeneration (AMD).

**Intervention(s)/ Methods:** Clinical imaging comprised serial infrared reflectance, eye-tracked spectral-domain optical coherence tomography (OCT), OCT angiography, and fluorescein angiography. Eye tracking, applied to the two preserved donor eyes, enabled correlation of clinical imaging signatures with high-resolution histology and transmission electron microscopy.

**Main Outcome(s) and Measure(s):** Histologic/ ultrastructural descriptions and diameters of vessels seen in clinical imaging.

**Results:** Six vascular lesions were histologically confirmed (type 3 MNV, n=3; deep retinal age-related microvascular anomalies (DRAMA), n=3). Pyramidal (n=2) or tangled (n=1) morphologies of type 3 MNV originated at the deep capillary plexus (DCP) and extended posteriorly to approach without penetrating persistent basal laminar deposit. They did not enter the sub-retinal pigment epithelium (RPE)-basal laminar space or cross Bruch’s membrane. Choroidal contributions were not found. The neovascular complexes included pericytes and non-fenestrated endothelial cells, within a collagenous sheath covered by dysmorphic RPE cells. DRAMA lesions extended posteriorly from the DCP into the Henle fiber and the outer nuclear layers, without evidence of atrophy, exudation, or anti-VEGF responsiveness. Two DRAMA lacked collagenous sheaths. External and internal diameters of type 3 MNV and DRAMA vessels were larger than comparison vessels in the index eyes and in aged normal and intermediate AMD eyes.

**Conclusions:** Type 3 MNV vessels reflect specializations of source capillaries and persist during anti-VEGF therapy. The collagenous sheath of type 3 MNV lesions may provide structural stabilization. If so, vascular characteristics may be useful in disease monitoring in addition to fluid and flow signal detection. Further investigation with longitudinal imaging before exudation onset will help determine if DRAMA are part of the type 3 MNV progression sequence.

## Introduction

Type 3 macular neovascularization (type 3 MNV) is a subtype of neovascular age-related macular degeneration (AMD).^1^ Unlike type 1 MNV, which arises from the choroid, type 3 MNV originates in the neurosensory retina.^2, 3^ Female gender, older age, and presence of subretinal drusenoid deposits confer risk for type 3 MNV.^4, 5^ Type 3 MNV is diagnosed in a third of Caucasian patients presenting with unilateral neovascular AMD^6^ and may be underestimated overall. Fellow eyes often convert to neovascular AMD within 3 years.^7-9^ Early lesions respond well to intravitreal anti-vascular endothelial growth factor (VEGF) therapy, unlike chronic lesions.^2, 10^ New information about type 3 MNV and related vascular anomalies from histopathology, as provided herein, could support improved detection and treatment decisions in affected patients.

Recent clinical imaging studies have elucidated type 3 MNV pathophysiology. Three stages are defined based on structural optical coherence tomography (OCT). Precursors to Stage 1 are hyperreflective foci (HRF) at the level of the deep capillary plexus (DCP), often near drusen. Stage 1 includes an intraretinal hyperreflective lesion and cystoid macular edema. In Stage 2, outer retinal disruption appears. At Stage 3, the hyperreflective lesion extends into the sub-RPE basal lamina (BL) space, associated with a pigment epithelium detachment.^2^ By color fundus photography, fluorescein angiography (FA), and OCT, type 3 MNV exhibit a specific regional distribution and pattern of hemorrhage.^11-13^ Lesions localize preferentially to the inner ring of the Early Treatment of Diabetic Retinopathy Study (ETDRS) grid. Flame-shaped intraretinal hemorrhages are located over type 3 MNV lesions, pointing toward the fovea. OCT angiography (OCTA) with 3-dimensional reconstruction and display shows vertically oriented components of type 3 MNV. Using this technology, Borelli et al describe two morphologic phenotypes (‘filiform’ and ‘saccular’) of advanced type 3 MNV.^14^

Prior clinicopathological correlation has elucidated some cellular detail corresponding to the OCT-based stages.^15, 16^ Neovascular complexes originating at the DCP have a vertical and downward trajectory. These complexes expand posteriorly and cross persistent basal laminar deposit (BLamD) to enter the sub-RPE-BL space. Some complexes appear like base-down pyramids that are ensheathed by collagenous material.^3, 16^ Participatory cells include macrophages, VEGF-positive fibroblasts, lymphocytes, Müller cell processes, and subducted RPE cells.^15, 16^ Hence, ischemia and inflammation may promote the development and progression of type 3 MNV.^15, 16^

Microvascular abnormalities involving the DCP include microaneurysms, telangiectasia, perifoveal exudative anomalous vascular complex (PEVAC), and capillary macroaneurysms. ^17-22^ PEVAC was initially described in non-AMD eyes as an isolated aneurysmal dilation of a retinal capillary originating between the superficial and deep plexuses, with exudation that is unresponsive to anti-VEGF.^17, 22, 23^ Deep retinal age-related microvascular anomalies (DRAMA) are recently proposed as DCP alterations in the setting of AMD findings like soft drusen and intraretinal HRF. Eyes with DRAMA show abnormal horizontal or vertical vessels with a diameter of >50 µm and/or a location below the posterior border of the outer plexiform layer (OPL).^24^ In contrast to type 1 MNV,^25^ precursors, early stages, and potential masqueraders for type 3 MNV, which may include DRAMA, are not described at the histologic level.^26^ Human eyes with longitudinal clinical imaging are especially valuable sources for such information.^3^

Herein we directly compared longitudinal OCT and angiographic signatures of intraretinal neovascularization and microvascular anomalies to corresponding histology. We analyzed both eyes of a single patient who had received intravitreal anti-VEGF treatments for type 3 MNV over the course of 5 years (right eye) and 9 months (left eye).

## Methods

### Compliance

Approval for this study was obtained by Institutional review at the University of Alabama at Birmingham (protocol #300004907). The study was conducted in accordance with the tenets of the Declaration of Helsinki and the Health Insurance Portability and Accountability Act of 1996.^27, 28^

### Clinical course

A white, pseudophakic woman in her 90’s received comprehensive ophthalmologic examination and multimodal imaging during a 5-year follow-up for bilateral type 3 MNV secondary to AMD. The patient presented 5 years prior to death with exudative type 3 MNV in the right eye. Over 5 years, she received a total of 37 intravitreal anti-VEGF injections over approximately 6 fluid resorption cycles in the right eye (12 × 0.5 mg/ 0.05 ml ranibizumab then 25 × 2 mg/ 0.05 ml aflibercept). One fluid resorption cycle is defined as the time in weeks and the number of injections needed from initial detection of intraretinal/ subretinal edema on OCT until complete absence of edema on OCT. The left eye was diagnosed with exudative type 3 MNV 4 years after the right eye. Over 9 months, the left eye received a total of 6 intravitreal anti-VEGF injections over approximately 2 fluid resorption cycles (12 × 0.5 mg/ 0.05 ml ranibizumab). Her general medical history included dyslipidemia and paroxysmal atrial fibrillation. Six months before death, the patient was diagnosed with gallbladder adenocarcinoma. Her last anti-VEGF treatment before death due to adenocarcinoma was 3 and 2 months for the left and right eye, respectively.

### Clinical image capture and analysis

All images were acquired using Spectralis HRA+OCT (Heidelberg Engineering, Heidelberg, Germany). Available for review were 11 (right eye) and 7 (left eye) eye-tracked spectral domain OCT volumes (6 mm x 6 mm horizontal and radial scans; 20° x 20° field). FA was available for both eyes at first presentation and 4 years later. One eye-tracked spectral domain OCTA volume (3 mm x 3 mm horizontal scans, 256 B-scans at 6 µm spacing, 10° x 10° field, ART 5, quality 34 dB) was obtained of the right eye 3.5 years after presentation.^29^ An investigational version of Heidelberg Eye Explorer (v. 6.16.100.701, Heidelberg Engineering, Heidelberg, Germany) was used for analysis, processing, and post-processing of data.^3,30^ Projection artifact was removed via 3-dimensional vessel-shape estimation and a Gaussian blur filter.^3, 30^ Raw (floating point) data were exported as a .VOL file.

Volume rendering enhances visualization of type 3 MNV, which is vertically oriented, and allows afferent/ efferent vascular connections to be identified, particularly in deeper retinal layers.^3, 31^ To visualize lesions at different angles of rotation, OCTA B-scans were first processed using linear quadratic estimation (noise variance estimate of 0.05 and a gain of 0.8; MATLAB version R2019b, Natick, Massachusetts: The MathWorks Inc.; 2019), followed by volume rendering and analysis (Imaris v9.5, Bitplane, Andor Technology plc.).^32^ The Filament Tracer tool was used to trace superficial arteries and veins after evaluation of dye circulation in FA. Video recording and still images were annotated to highlight structural and flow details.

Previous reports indicated both eccentricity- and hemifield-dependent asymmetries in the spatial distribution of type 3 MNV.^11, 12, 33^ Thus, for potential mechanistic insight into tissue-level associations, location of vascular lesions was documented. Lesion distance from the fovea was calculated using the OCT volume and NIR en face image using a custom ImageJ plug-in ‘Spectralis Browser OCT’, available at https://sites.imagej.net/CreativeComputation/. Meridional position was documented using the sectors of the ETDRS grid.^34,35^

### Histology preparation and image analysis

As described, ^36^ globes were recovered 2:05 hours after death and preserved in buffered 1% paraformaldehyde and 2.5% glutaraldehyde. Pre-mortem eye-tracked OCT volumes were registered to post-mortem OCT volumes of the same globes.^37^ For high-resolution histology over large areas, a rectangular tissue block containing fovea and optic nerve was post-fixed in 1% osmium – tannic acid – paraphenylenediamine and embedded in epoxy resin. A tissue block 8 mm x12 mm wide was processed for stepped sections at 30-60 μm intervals. Interleaved 30 μm-thick slabs were re-embedded for transmission electron microscopy (TEM). Sub-micrometer sections stained with toluidine blue were scanned using a 60X oil immersion objective.^38^ Tissue sections on 112 (right eye) and 87 (left eye) glass slides spanning a distance of 5453 µm (right eye) and 4243 µm (left eye) centered on the two foveas were matched to clinical OCT scans by comparing overall tissue contours.

DRAMA vessels are defined by a diameter criterion (>50 µm),^24^ and our previous and current observations indicated that a collagenous sheath surrounds type 3 MNV neovessels. Further, OCTA shows only the moving blood cell column and not the collagenous sheath. Therefore, we manually measured internal (luminal) and external cross-sectional diameters of vessels (‘oval’ tool, FIJI Is Just; ImageJ 2.0.0-rc-69/1.52p; www.fiji.sc;). Type 3 MNV vessels meandered over several glass slides, and sections on each slide were measured. To contextualize neovessel measurements, DCP vessels on either side of the area directly involved in exudation in the index case were also measured. Further, vessels aligned along the outer border of the INL of intermediate AMD eyes and age-similar controls (n= 8 each) on the Project MACULA website of AMD histopathology were also measured.^25^ Because vessels could run longitudinally within a section, we report a minimal cross-section diameter (approximated by Feret diameter in ImageJ for ellipses). Due to small numbers, these data were not analyzed statistically.

## Results

**Table S1** lists all figures to provide an overview.

### Classification of lesions and longitudinal clinical imaging

We first classify vascular complexes in the two eyes at one time point, then describe them in longitudinal clinical imaging and detailed histology. **Table 2** lists hyperfluorescent lesions seen by FA 11 months before death (**Figure 1**). Six of 7 lesions localized to the inner ring of the ETDRS grid (0.5-1.5 mm eccentricity), with the seventh at 1.52 mm. There was no predilection for any one sector. Six of 7 lesions were confirmed as vascular by histology, and the seventh (**Figure S2**) could not be found. Lesion morphology was categorized as tangled type 3 MNV (n=1), pyramidal type 3 MNV (n=2), or DRAMA (n=3). As seen previously, ^3, 16^ pyramidal type 3 MNV was defined as a focal, vertically extending neovessel complex. Tangled type 3 MNV was defined as a horizontally extending neovessel complex. DRAMA was defined as anomalous vascular elements extending posterior to the DCP and into the HFL/ONL, i.e., anterior to the ELM.^24^ Hyperfluorescent lesions in the right eye due to window defects were reported elsewhere.^39^

**Table 2.** Spatial distribution and vascular lesion types in two eyes of the index case.

| Eye, lesion number,<br>color in Fig. 1 | ETDRS<br>ring & sector | Lesion type | intravitreal<br>anti-VEGF | Analysis |
| --- | --- | --- | --- | --- |
| OD 1 (green) | Inner, S | T3MNV, tangled | 37 | LM |
| OD 2 (yellow) | Inner, N | T3MNV, pyramidal | 37 | LM |
| OD 3 (fuchsia)** | Inner, T | DRAMA | 37 | LM |
| OS 1 (yellow) | Inner, S | DRAMA | 6 | LM |
| OS 2 (white) | Outer, I | not found | 6 | n.a. |
| OS 3 (fuchsia) | Inner, I | DRAMA | 6 | LM, EM |
| OS 4 (green) | Inner, N | T3MNV, pyramidal | 6 | LM, EM |

T3MNV, type 3 macular neovascularization; DRAMA, deep retinal age-related microvascular anomaly; LM, light microscopy; EM, electron microscopy. OD, right eye; OS, left eye.
ETDRS sectors (delimited by 45° lines): S, superior; I, inferior; N, nasal; T, temporal
Within each eye, lesions seen on fluorescein angiography were numbered clockwise, starting at 12:00 (ETDRS grid superior sector).
\* Span width of tangled vascular lesion: 520-769 µm; \*\* pair of vascular lesions.

**Figure 1.**
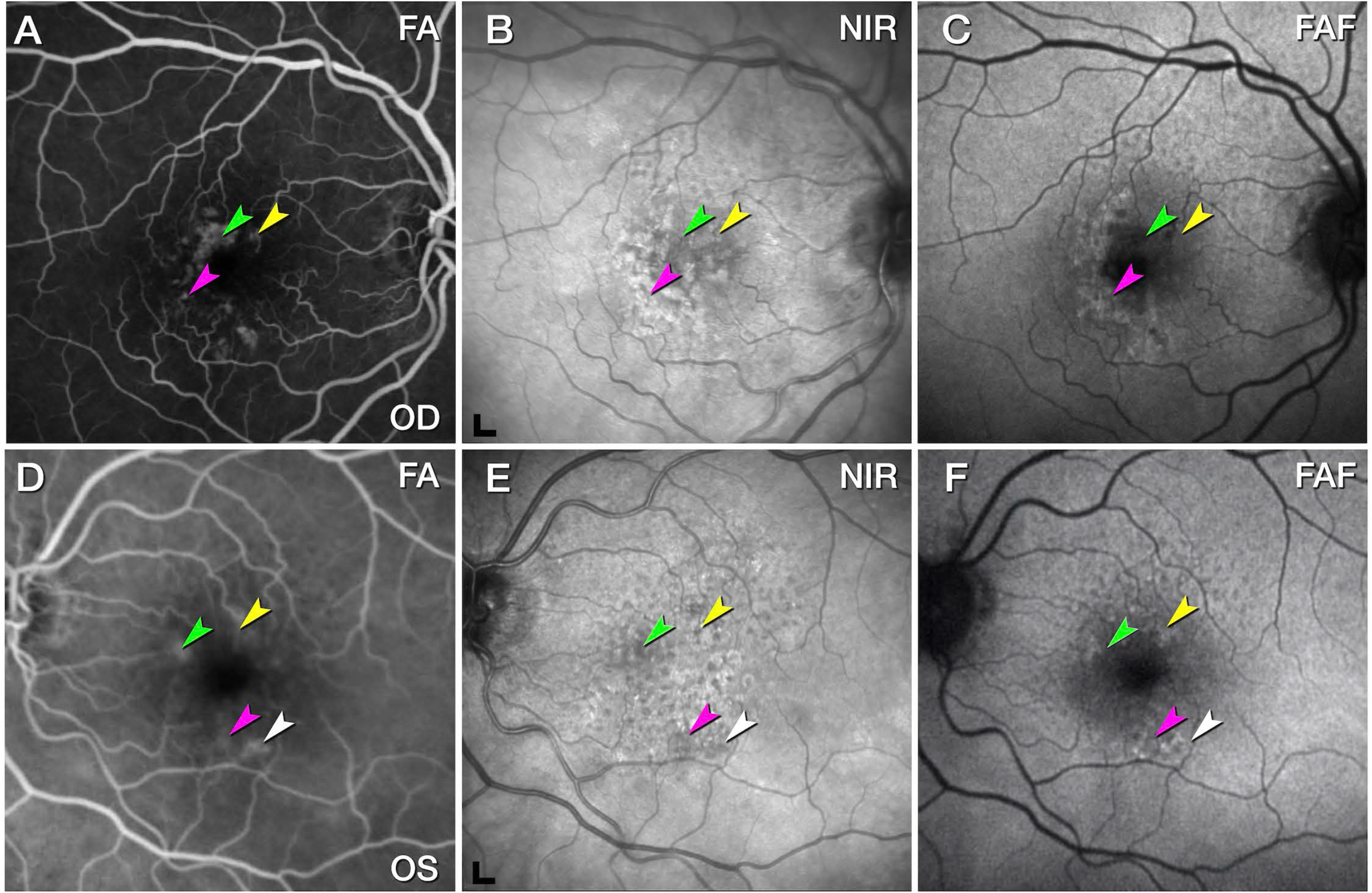
Multimodal retinal imaging of both eyes, 11 months before death. **A & D**. Fluorescein angiography (FA) in venous (**A**) and recirculation phases (**D**) shows multiple instances of hyperfluorescence. Vessels were found by histology at the green, yellow, and fuchsia arrowheads in the right (**A**), and left eye (**D**). At the white arrowhead in the left eye, no vessel could be found in histology. **B & E**. Near-infrared reflectance imaging (NIR) shows reduced reflectance in areas of angiographic leakage (arrowheads), possibly due to retinal edema. Soft drusen exhibit hypo- and hyperreflective mottling. Scale bar 200 µm. **C & F**. Fundus autofluorescence (FAF; λ_ex_= 488 nm) highlights subretinal drusenoid deposits especially superior to the fovea. Lesion number, type, and spatial distribution are listed in **Table 2**.

At initial presentation 5 years before death, the right eye exhibited multiple instances of MNV secondary to AMD with multifocal leakage on FA (**Figure S3**). At this time, fluid was not detected in the left eye (**Figure S4**). Four years later and 11 months before death (**Figure 1**), the left eye was diagnosed with exudative type 3 MNV due to AMD. By this time, the right eye had received 32 intravitreal anti-VEGF injections at intervals of 4-8 weeks resulting in 4 fluid resorption cycles.

### Pyramidal (OD 2, OS 4) and tangled (OD 1) type 3 MNV

Pyramidal type 3 MNV (OS 4) closely resembles previously described OD 2 ^4^ in terms of location, OCT appearance, and histologic features (**Figure 1, Figure 5, Figure 6**, respectively). In brief, OS 4 is a vertically oriented, pyramidal-shaped, intraretinal lesion located in the nasal sector of the ETDRS inner ring, with heterogeneous hyperreflectivity on OCT (**Figure 5**). On histology, this lesion corresponds to a collagen-ensheathed neovascular complex that extends from the OPL/INL border through the HFL /ONL (**Figure 6A**). The complex is flanked by RPE cells that extend off the top of the pyramid and along a DCP vessel (**Figure 6B**). The pyramid base adheres to denuded and persistent BLamD, which drapes a calcified druse (**Figure 6A&B**).^4^ The vessel did not enter the subRPE-BL space, and no choroidal contribution was found.

**Figure 5.**
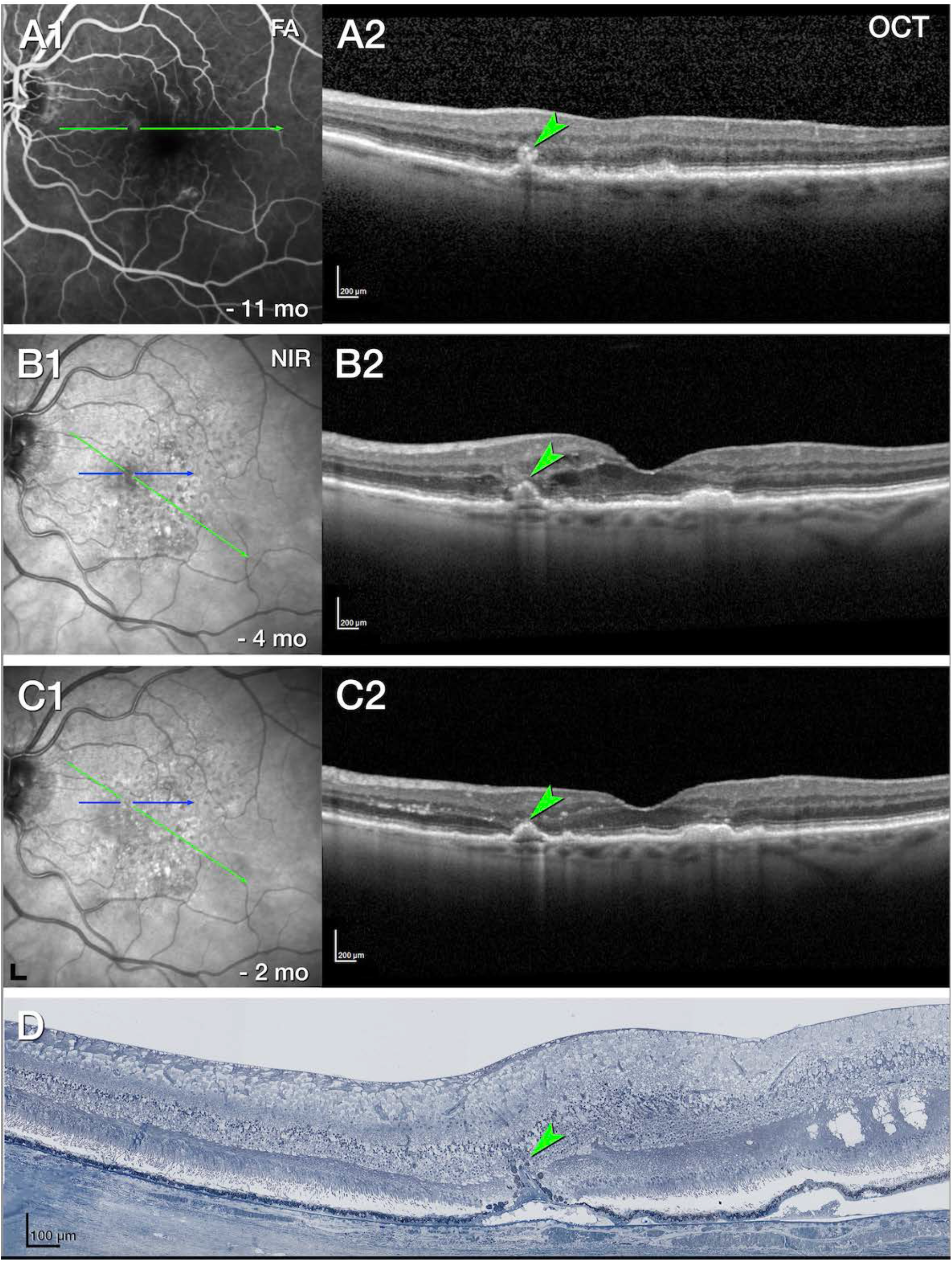
Multimodal imaging, clinical course, and histology of pyramidal type 3 MNV, OS 4. **A**. Venous phase fluorescein angiography (FA, **A1**) shows mild leakage at site of type 3 MNV 11 months before death. Green lines on FA represent optical coherence tomography (OCT) B-scans. Horizontal OCT B-scan (**A2**) displays a hyperreflective lesion, hyperreflective foci (HRF), and small intraretinal cysts. **B**. Blue lines on near-infrared reflectance (NIR, **B1**), represents histology section in **D**. Radial OCT B-scan (**B2**) displays enlarged intraretinal cysts, after 5 total injections and 8 weeks following the prior injection. There is also subsidence of the outer plexiform layer (OPL) and external limiting membrane (ELM). **C**. On radial OCT B-scan (**C2**), intraretinal fluid is reduced after 6 injections and 6 weeks following the prior injection. Numerous HRF are present in the inner nuclear layer (INL; **C2;** scale bar 200 µm). **D**. On histology, type 3 MNV is a pyramidal complex bounded by retinal pigment epithelium cells. It extends from the INL/ OPL border to basal laminar deposit draping a calcified druse. Structural damage to Henle’s fiber layer at the right of the panel (green asterisk) may indicate an area of prior intraretinal fluid. Magnified histology is shown in **Figure 6**. Scale bar 100 µm. Blue line, histology section. Time in months, time before death.

**Figure 6.**
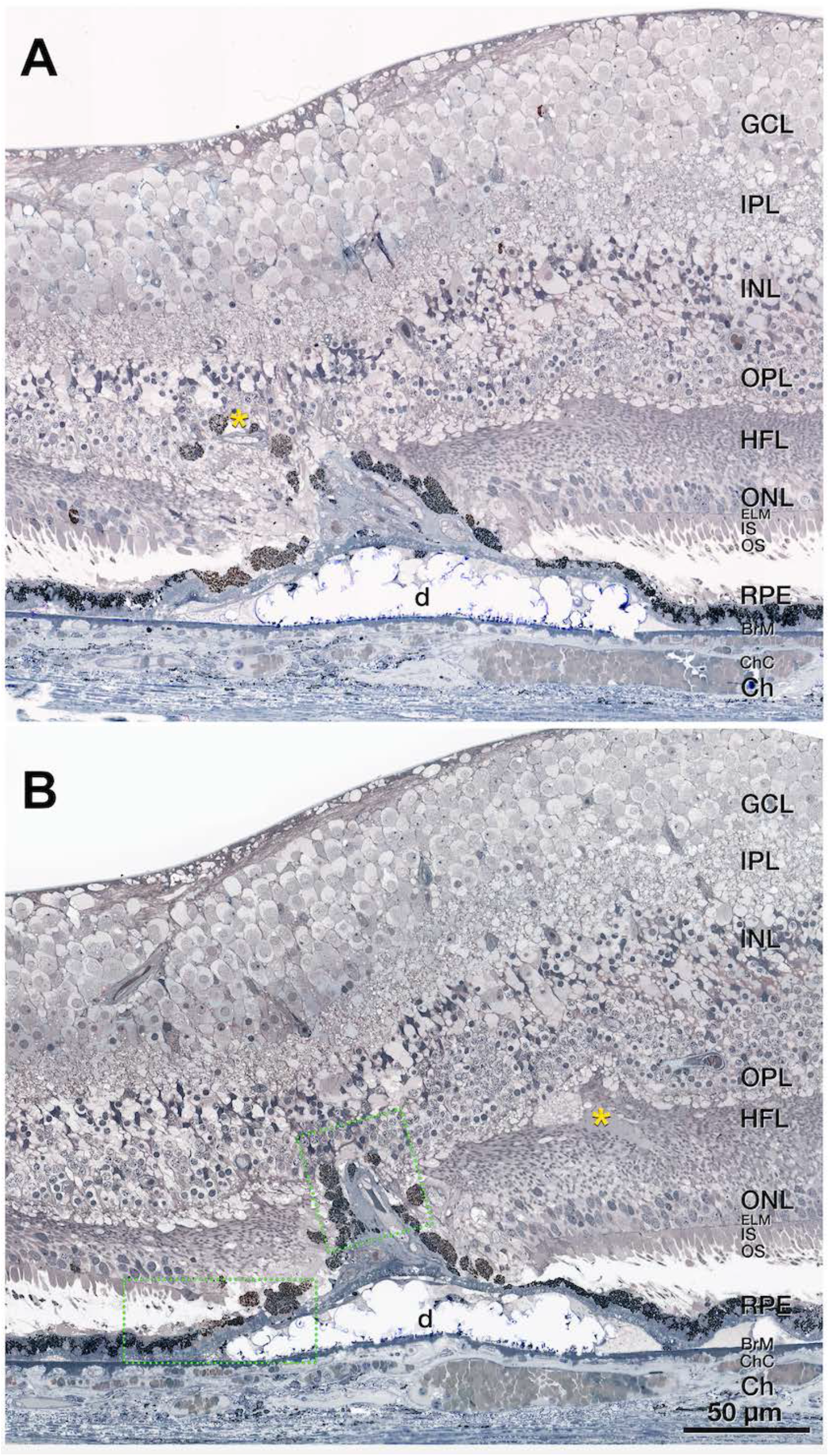
Pyramidal vascular complex in type 3 MNV, OS 4. **A, B**. A pyramidal complex includes neovessels ensheathed by thick layers of collagenous material and flanked by retinal pigment epithelium cells. The complex extends from the inner nuclear layer (INL) through the outer plexiform layer, Henle fiber layer, outer nuclear layer (OPL, HFL, ONL, respectively) and terminates at the basal laminar deposits draping a calcified druse. Bruch’s membrane appears intact with no evidence of a choroidal contribution to the neovessel complex. The external limiting membrane (ELM) descends at both sides of type 3 MNV base. There is some fluid at the OPL-HFL border (yellow asterisk in **B**). Areas within dotted green boxes are shown in **Figure 10**.

Transmission electron microscopy of this pyramidal lesion reveals endothelial cells and pericytes (**Figure 7A**). Endothelial cells are not fenestrated (**Figure 7B**), like endothelial cells of the DCP and unlike endothelial cells of the choriocapillaris (**Figure 7C&D**). Other vessel wall components, e.g., smooth muscle cells, connective tissue, or a 3-layer arterial configuration, cannot be identified. In RPE cells arrayed along the sloping sides of the calcified druse, lipofuscin becomes less electron-dense, smaller, and less tightly packed (**Figure 7E**), in a smooth transition. These findings suggest transdifferentiation, rather than ingestion of RPE organelles by invading phagocytes.

**Figure 7.**
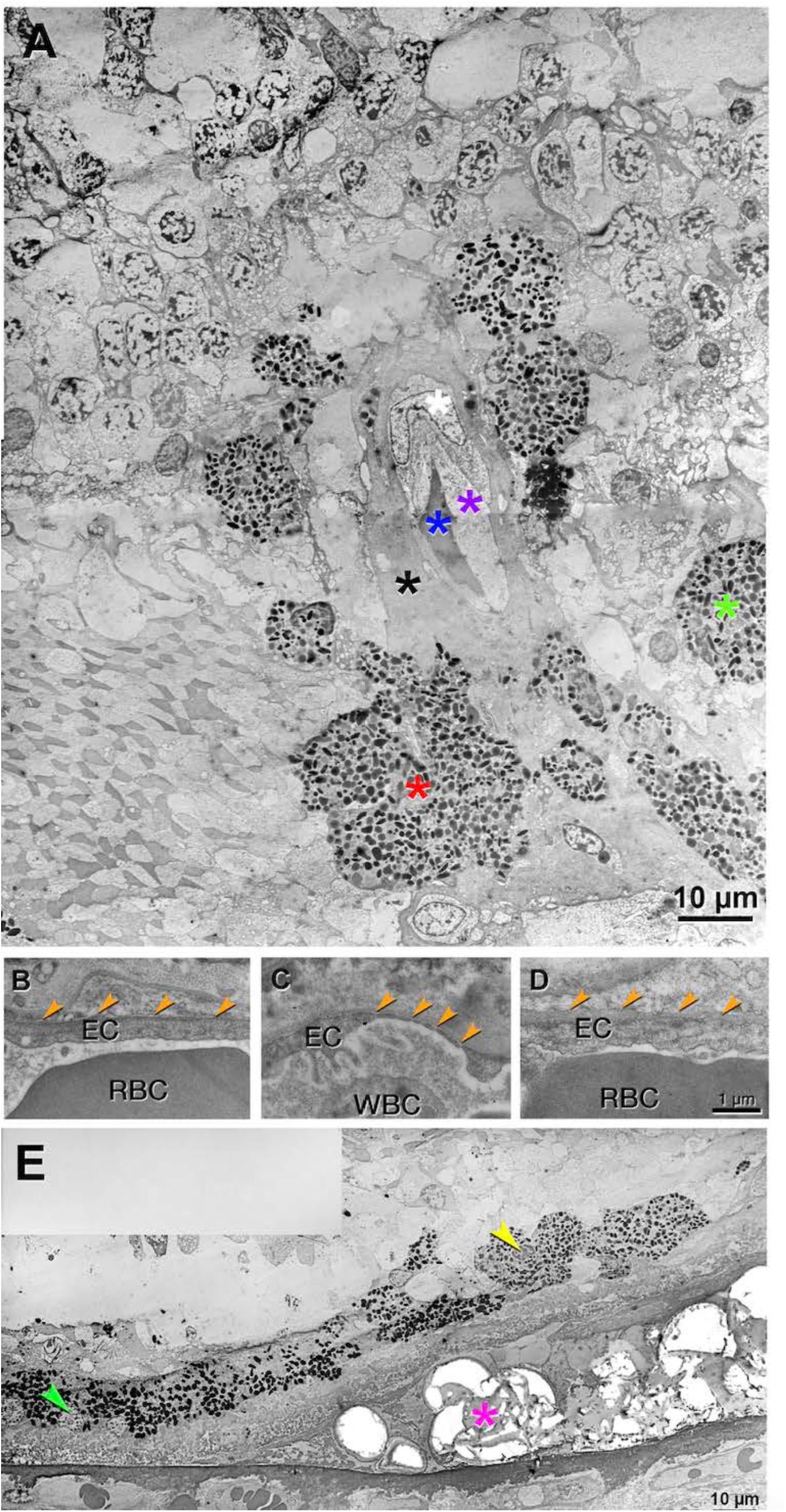
Transmission electron microscopy of pyramidal type 3 MNV, OS 4. See Figure 8 for light microscopy of OS4. **A**. Neovessel with erythrocyte in the lumen (blue asterisk) is ensheathed by endothelium, pericyte, and collagenous material (purple, white, and black asterisks, respectively). Surrounding retinal pigment epithelium (RPE) cells merge into multi-nucleated cells (red asterisk) or disperse into the Henle fiber layer (HFL, green asterisk). Phagolysosomes are not visible in the RPE cells. **B-D**. Endothelial cells in the neovessel and comparison vessels are displayed. The lumen is located at the bottom of all panels. **B**. No fenestrations are detected in the neovessel (orange arrowheads). **C**. Fenestrations are visible in the choriocapillaris (orange arrowheads). **D**. No fenestrations are detected in the deep capillary plexus (orange arrowheads). **E**. Atop the calcified druse (fuchsia asterisk), RPE lipofuscin is more electron-dense at the druse base than at the druse top (left vs right in the panel). These changes are consistent with transdifferentiation.

Next we consider tangled type 3 MNV (OD 1), for which en face OCTA (**Figure 8A2**) and OCT with flow overlay (**Figure 8A3**) shows persistent flow signal. Over time, the extent of intraretinal fluid surrounding OD 1 in the INL and HFL fluctuates (**Figure 8B2, Figure 8C2**, respectively). Volume rendering of structural OCT and OCTA together highlights flow within a hyperreflective lesion at the ONL and bacillary layer (**Figure S9A, Video S1**). Inflow and outflow vessels can be connected to a superficial artery and vein, respectively (**Figure S9B&C**). The RPE/BrM complex below the lesion is split by hyporeflective material producing a double layer sign (**Figure 7B2**).^40^ Numerous HRF are present in the INL (**Figure 8C2**).

**Figure 8.**
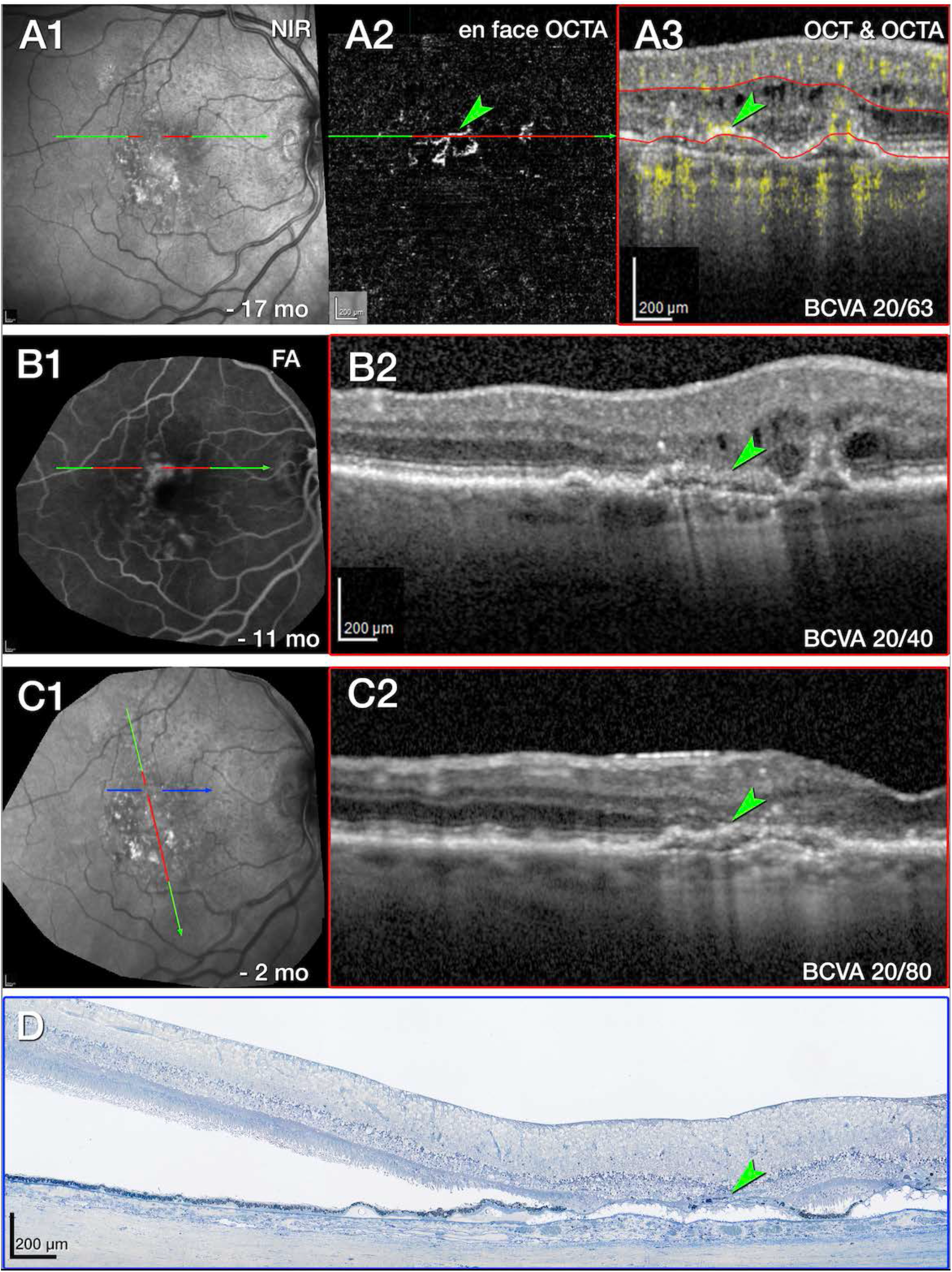
Multimodal imaging, clinical course, and histology of tangled type 3 MNV, OD 1. **A**. Green and red lines on near-infrared reflectance (NIR, **A1**) represent optical coherence tomography (OCT) B-scans corresponding to en face OCT angiography (OCTA, **A2**) and OCT B-scan with flow signal overlay (**A3**). After 29 total injections and 8 weeks following the prior injection (**A2&A3**), flow signal persists within the tangled hyperreflective type 3 MNV lesion (green arrowhead). The RPE/Bruch’s membrane complex is split by hyporeflective material and appears as a “double layer” sign without flow signal (**A3**). Red lines in **A2** indicate the segmentation boundaries [outer plexiform layer (OPL)-retinal pigment epithelium (RPE)] used to create the en face OCTA (**A2**). **B**. Fluorescein angiography (FA, **B1**) shows late venous phase hyperfluorescence, after 32 total injections and 8 weeks following the last injection. OCT B-scan (**B2**) shows intraretinal fluid in the inner nuclear and Henle fiber layers surrounding the type 3 MNV lesion (green arrowhead). **C**. NIR (**C1**) and radially oriented OCT B-scan (**C2**) shows intraretinal fluid adjacent to the tangled type 3 MNV lesion (green arrowhead), after 36 total injections and 8 weeks following the prior injection. Blue line in **C1** represents histology section in panel **D.** **D**. Panoramic histology shows a horizontally oriented tangled type 3 MNV lesion (green arrowhead), partly bounded by RPE cells (scale bar 200 µm). The complex extends from the inner nuclear layer border to a chipped out calcified druse, which correlates to the double layer sign in B1. The druse is draped by basal laminar deposit and lacks RPE at its apex. Time in months, time before death; BCVA, best corrected visual acuity.

Histologic analysis reveals components of tangled type 3 MNV (OD 1, **Figure 8D**). Magnified histology shows a vascular complex spanning 249 µm horizontally, extending into the superior perifovea (**Figure S10**). The complex is partly ensheathed by collagenous material and flanked by intraretinal RPE cells. The INL/OPL border subsides where the vascular lesion extends through the HFL/ONL (**Figure S10C**). The ELM subsides at both edges of the calcified druse (yellow arrows in **Figure S10B**). Bruch’s membrane appears intact without evidence of a choroidal contribution to the MNV lesions, or evidence of MNV contributing to the OCT double layer sign (**Figure S10A-C**). Neovessels within the INL are moderately dilated, suggesting drainage venules (**Figure S10A**).^41^

To summarize, vascular geometry and thickness of the collagenous sheath differentiates tangled versus pyramidal type 3 MNV. The sheath surrounding the tangled complex is thin (**Figure S10**), and that surrounding pyramidal type 3 MNV is thick (**Figure 6**). Otherwise, the two subtypes are comparable.

### DRAMA, in the setting of anti-VEGF therapy

As shown below, all three instances of DRAMA (OS 1, OS 3, and OD 3) exhibit mild hyperfluorescence on venous and recirculation phase FA 11 months before death. In histology, all three localize above a horizontal ELM, signifying lack of atrophy.

OS 1 includes a vessel extending from the INL into the ONL (**Figure 11A**). Like type 3 MNV lesions, this DRAMA lesion is located above a soft druse with BLamD and altered RPE at its apex (**Figure 11C**). Unlike type 3 MNV, OS 1 does not have a collagenous sheath, and the OPL does not subside (**Figure 11C2**).

**Figure 9.**
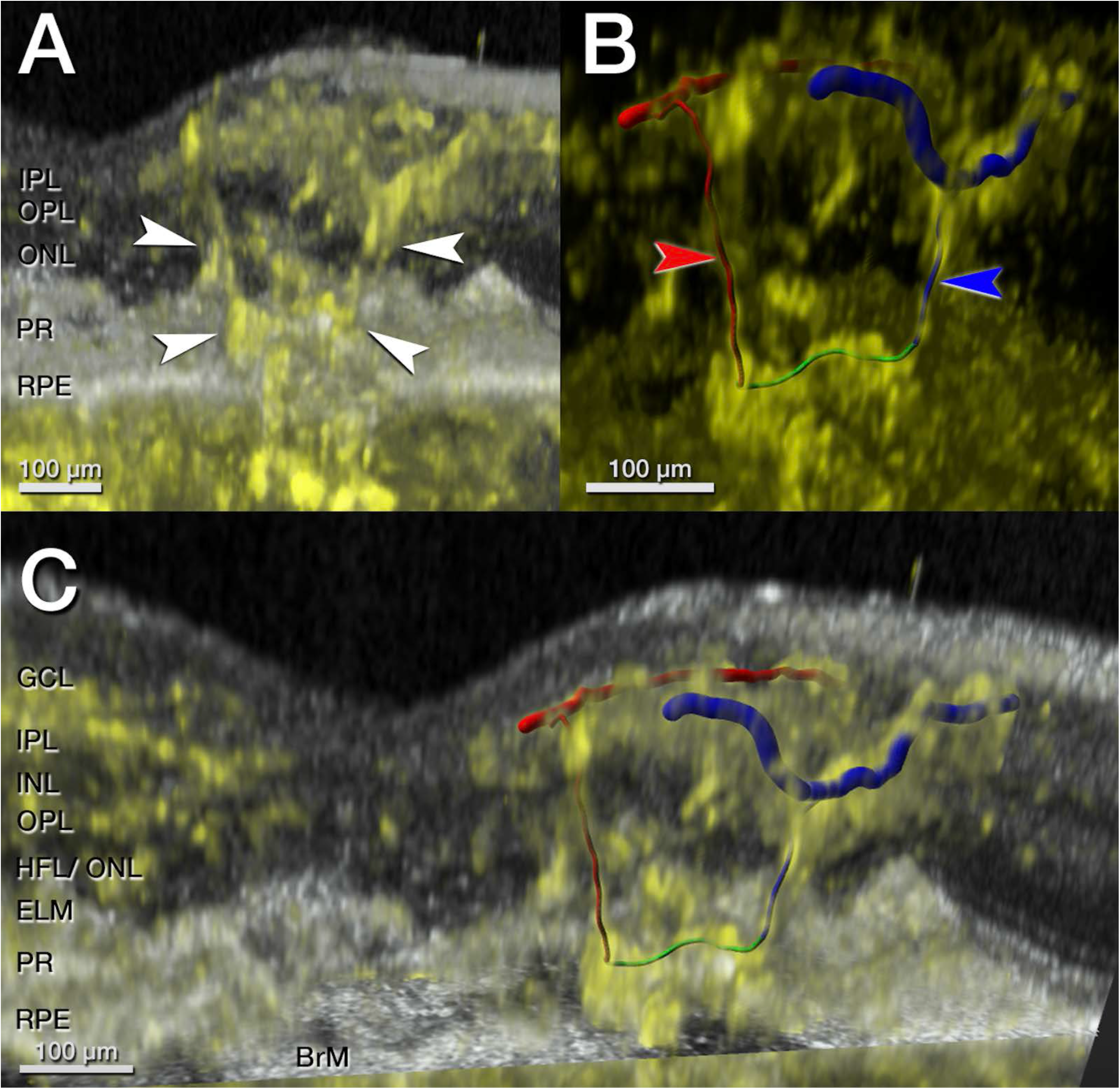
Volume rendering of structural optical coherence tomography (OCT, gray) and OCT angiography (OCTA, yellow) of tangled type 3 MNV, OD 1. **A**. Neovascular blood flow within a hyperreflective structure (white arrowheads) is observed at the level of the outer nuclear layer (ONL). **B**. Three-dimensional analysis of neovascular blood flow depict an anastomosis above the retinal pigment epithelium (RPE)/ Bruch’s membrane (BrM; green section) and a tangled structure connecting to the superficial artery (red arrow) and vein (blue arrow). **C**. Volume rendering of OCTA with orthogonal structural sections evidence a tangled neovascular lesion.

**Figure 11.**
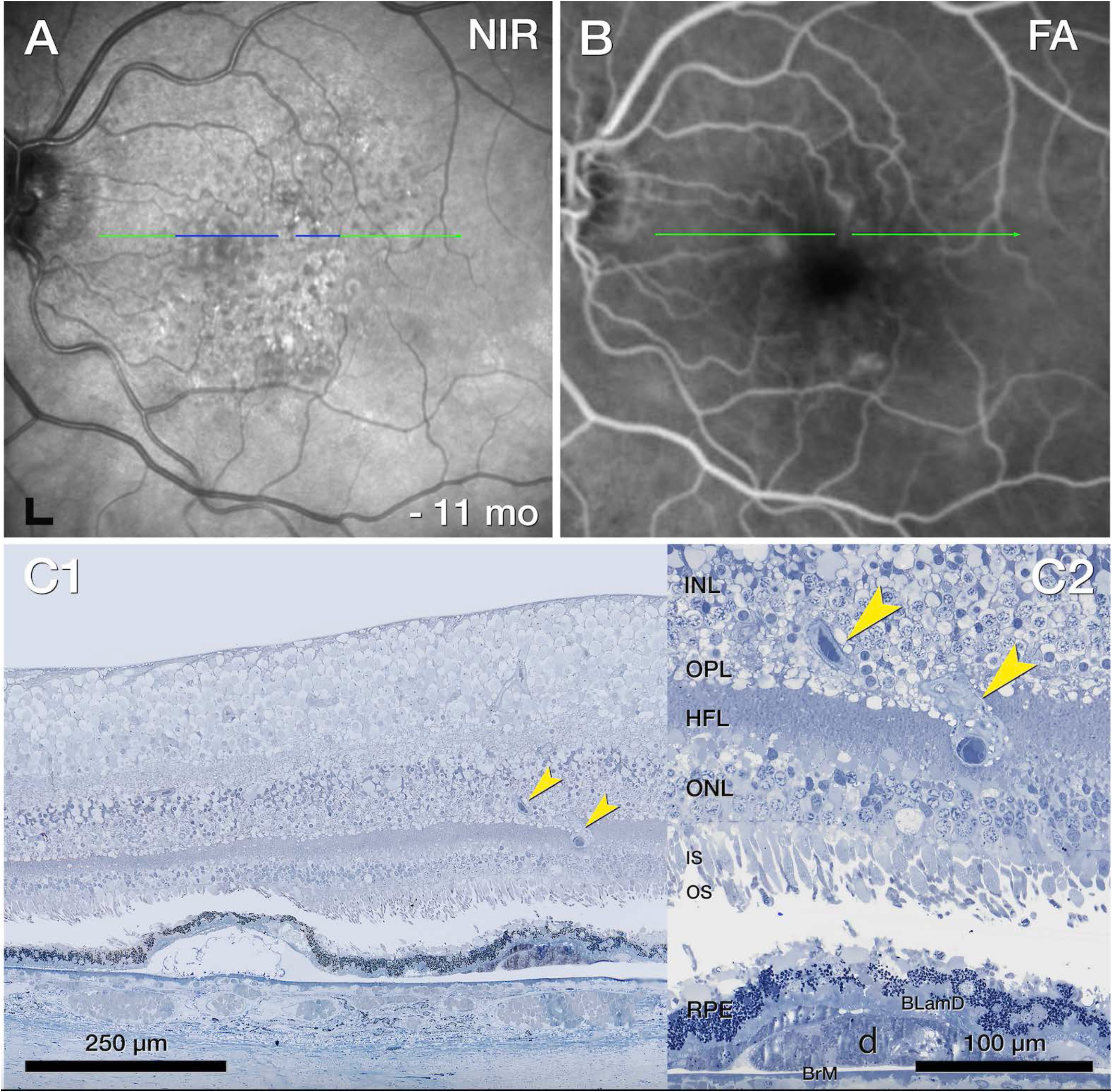
Deep retinal age-related microvascular anomaly (DRAMA), OS1. **A**. Near infrared reflectance (NIR) shows drusen and subretinal drusenoid deposits 11 months before death. Blue line, plane of histology section. **B**. Fluorescein angiography (FA) recirculation phase shows minimal leakage at the site of DRAMA. **C**. On histology, a vessel extends downwards (yellow arrowheads) from the inner nuclear layer (INL) into the outer plexiform layer (OPL) above a druse.

Corresponding to FA, OCT of OS 3 shows an intraretinal hyperreflective, stacked lesion, without intraretinal cysts (**Figure 12A**). Over time, a plume of HRF extends nasally, still without evidence of cysts (**Figure 12C**).^42, 43^ In histology, an RPE tower atop a soft druse extends into the OPL (**Figure 12D**). This RPE complex surrounds like a gripping hand a vessel extending downward from the DCP (**Figure 13A&B**). By light microscopy, the extending vessel in OS 3 resembles DRAMA OS 1 (**Figure 11**) in its location above a soft druse with thick BLamD and a thinned RPE layer. By transmission electron microscopy, OS 3 (**Figure S14**) resembles type 3 MNV lesions but lacks a collagenous sheath. Endothelial cells in OS 3 lack fenestrations (**Figure 13C**), like endothelial cells of the DCP and unlike endothelial cells of the choriocapillaris (**Figure 13D&E**) Other vessel wall components cannot be identified. The RPE complex is multicellular, with some multinucleated cells (**Figure S14**). Organelle packing and electron density is similar to in-layer RPE cells (not shown).

**Figure 12.**
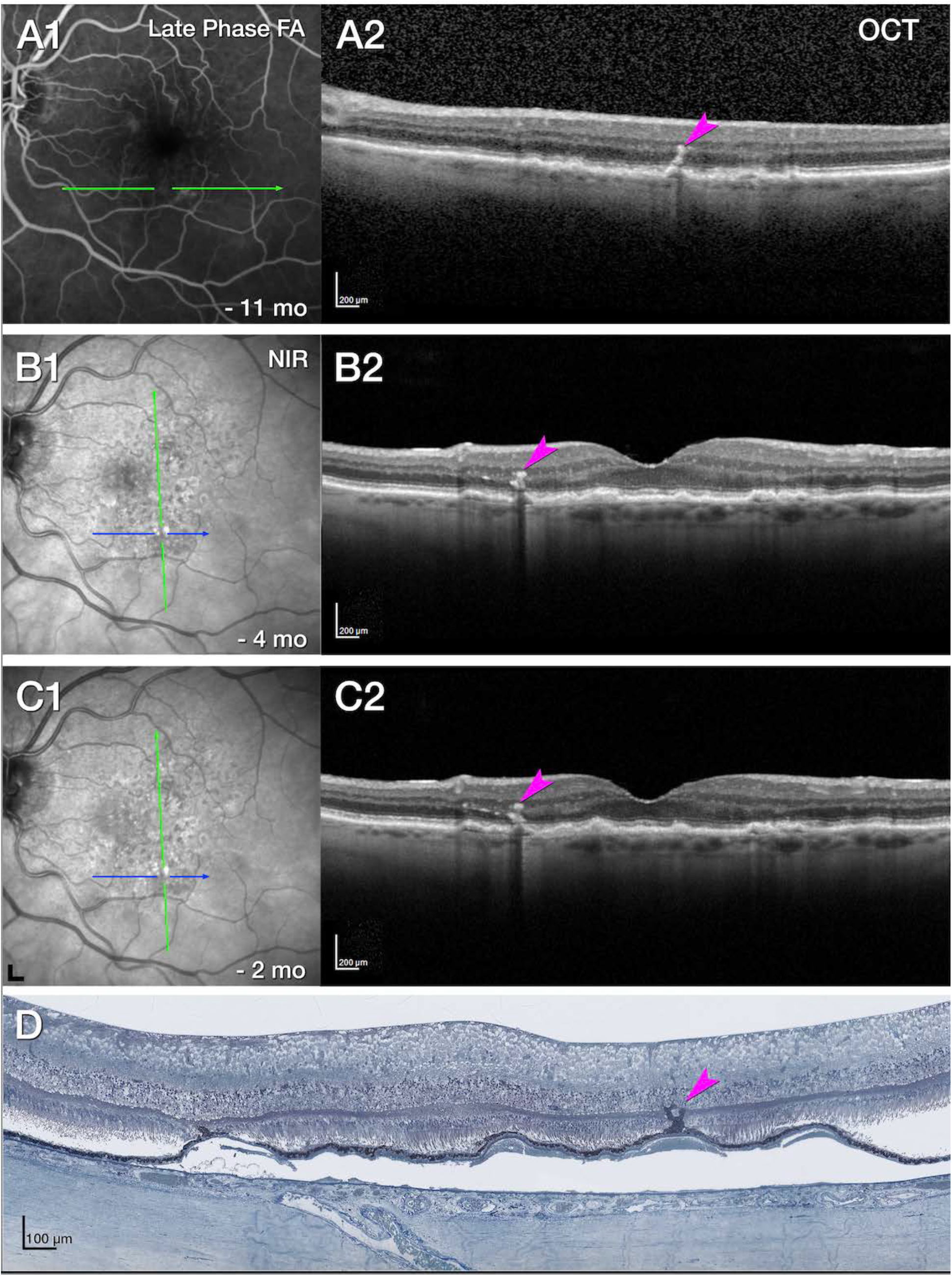
Multimodal imaging and histology of deep retinal age-related microvascular anomaly (DRAMA) with intraretinal RPE complex, OS 3. **A**. Fluorescein angiography (FA) venous phase shows faint staining, 11 months before death (**A1**). Optical coherence tomography (OCT, **A2**) shows a stack of intraretinal hyperreflective foci (HRF, fuchsia arrowhead). No intraretinal cysts are visible. **B**. After 6 total injections and 6 weeks following the prior injection, the stacked lesion (fuchsia arrowhead) is stable, without cysts. A plume of HRF extend nasally 2 months before death. Green lines, OCT B-scans; blue line, histology section. **C**. On histology, a retinal pigment epithelium tower (fuchsia arrowhead) rises upward from a soft druse. Cells surround a vessel and extend into the outer plexiform layer. Magnified histology is shown in **Figure 13**.

**Figure 13.**
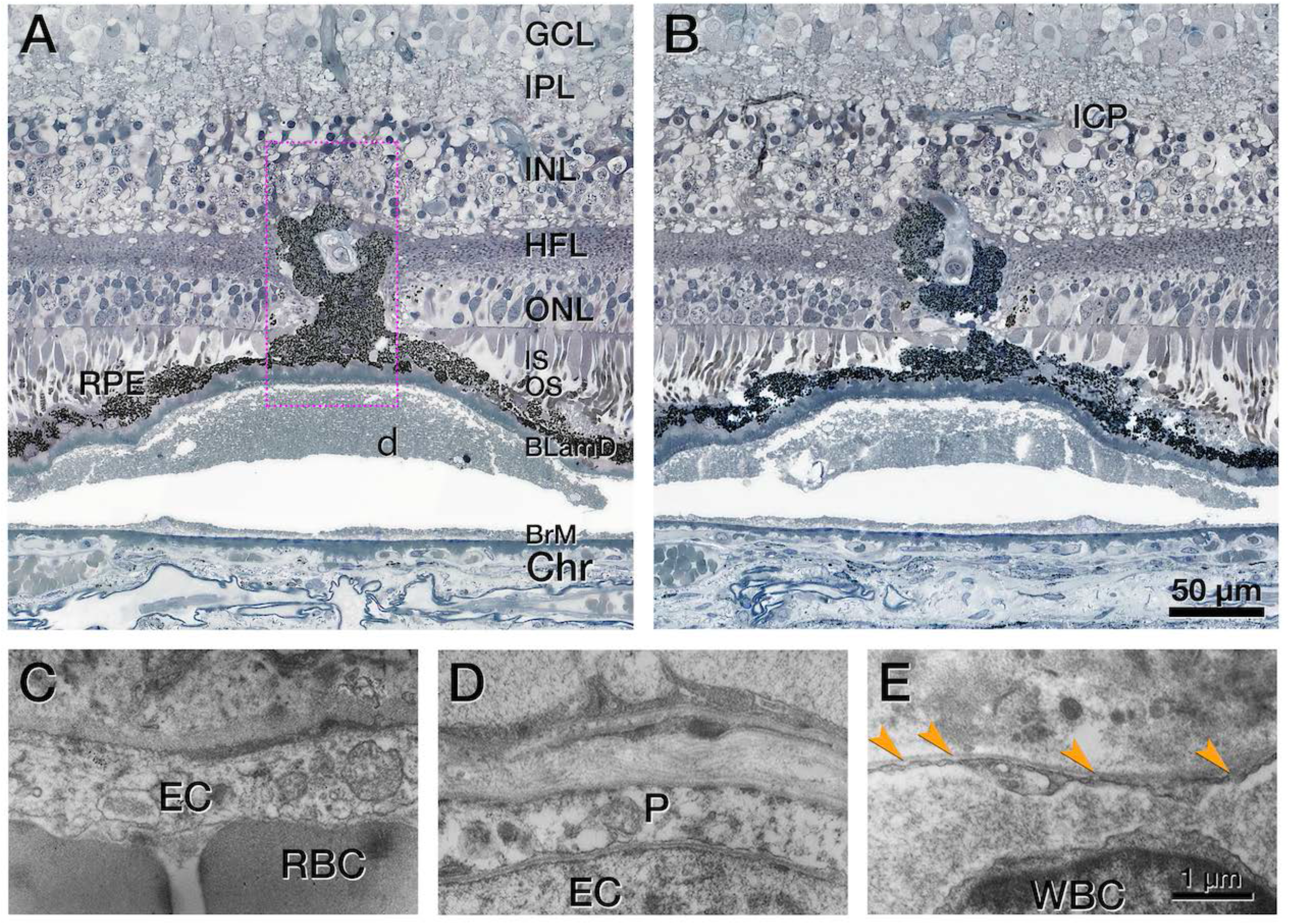
RPE complex associated with deep retinal age-related microvascular anomaly (DRAMA), OS 3. **A**. A “hand” shaped complex consisting of retinal pigment epithelium (RPE) cells surrounds vessel extending downward from the deep capillary plexus. Electron microscopy of the area in the dotted fuchsia box is shown in **Figure S14.** **B**. The RPE complex extends upward to the inner nuclear layer (INL). On either side of this complex, there is no subsidence of outer plexiform layer (OPL) or external limiting membrane (ELM), and the outer nuclear layer (ONL) is thinned. The complex emanates from a continuous RPE layer over basal laminar deposit (BLamD) and a soft druse. The druse is artifactually detached from Bruch’s membrane (BrM). **C-E**. Endothelial cell (EC) ultrastructure in DRAMA and comparison vessels are shown. Vascular lumen is at the bottom of all panels. **C**. No fenestrations are visible in the DRAMA (red blood cell, RBC). **D**. No fenestrations are visible in the deep capillary plexus (pericyte, P). **E**. Fenestrations are visible in the choriocapillaris endothelium (orange arrowheads; white blood cell, WBC).

OCTA flow overlay of DRAMA OD 3 shows an intraretinal hyperreflective lesion containing a pair of vascular outpouchings (**Figure 15C**). Corresponding histology displays a pair of deeply descending vessels without significant intraretinal fluid (**Figure 15D**). Like type 3 MNV, the vessel complex of this DRAMA is ensheathed by a thin layer of collagenous material and dives from the INL into the HFL (**Figure 16A**). Unlike type 3 MNV, the vessels do not extend past the HFL, and there is no subsidence of the ELM. RPE organelles appear near the vessel complex (**Figure 16B**).

**Figure 15.**
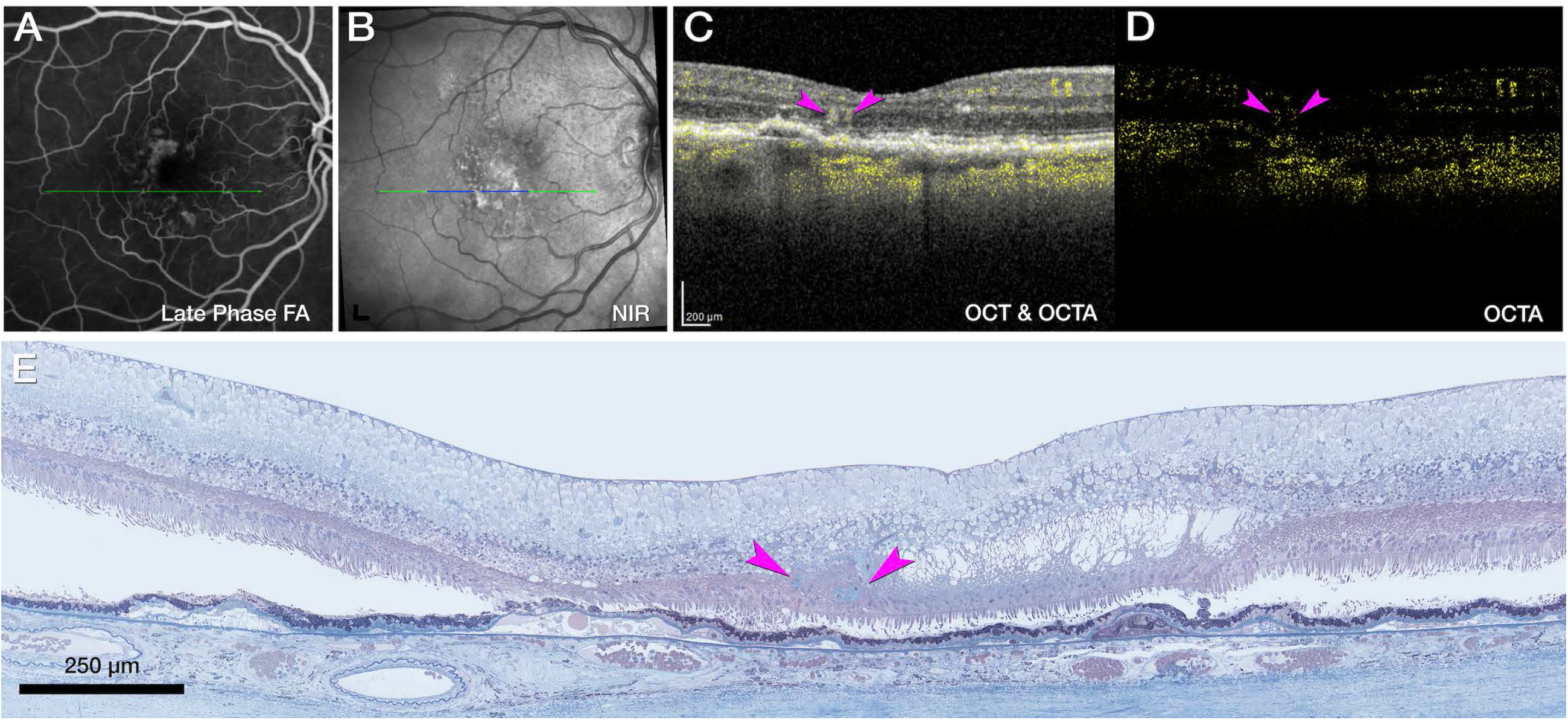
Multimodal imaging of deep retinal age-related microvascular anomalies (DRAMAs), OD 3. **A**. Fluorescein angiography (FA) venous phase shows mild hyperfluorescence 11 months before death. **B**. Green and blue lines on near-infrared reflectance (NIR), represent optical coherence tomography (OCT) B-scan (C) and histology section (D**). C**. Horizontally oriented OCT B-scan with OCT angiography (OCTA) flow overlay shows cyst-like spaces in the Henle fiber layer adjacent to a pair of hyperreflective DRAMAs (fuchsia arrowheads). The retinal pigment epithelium/Bruch’s membrane complex to the left is split by hyporeflective material. **D**. OCTA shows a pair of intraretinal flow signals (fuchsia arrowheads). **E**. Histology shows a pair of vessels extending from the inner nuclear layer border to the outer nuclear layer. Magnified histology is shown in **Figure 16**.

**Figure 16.**
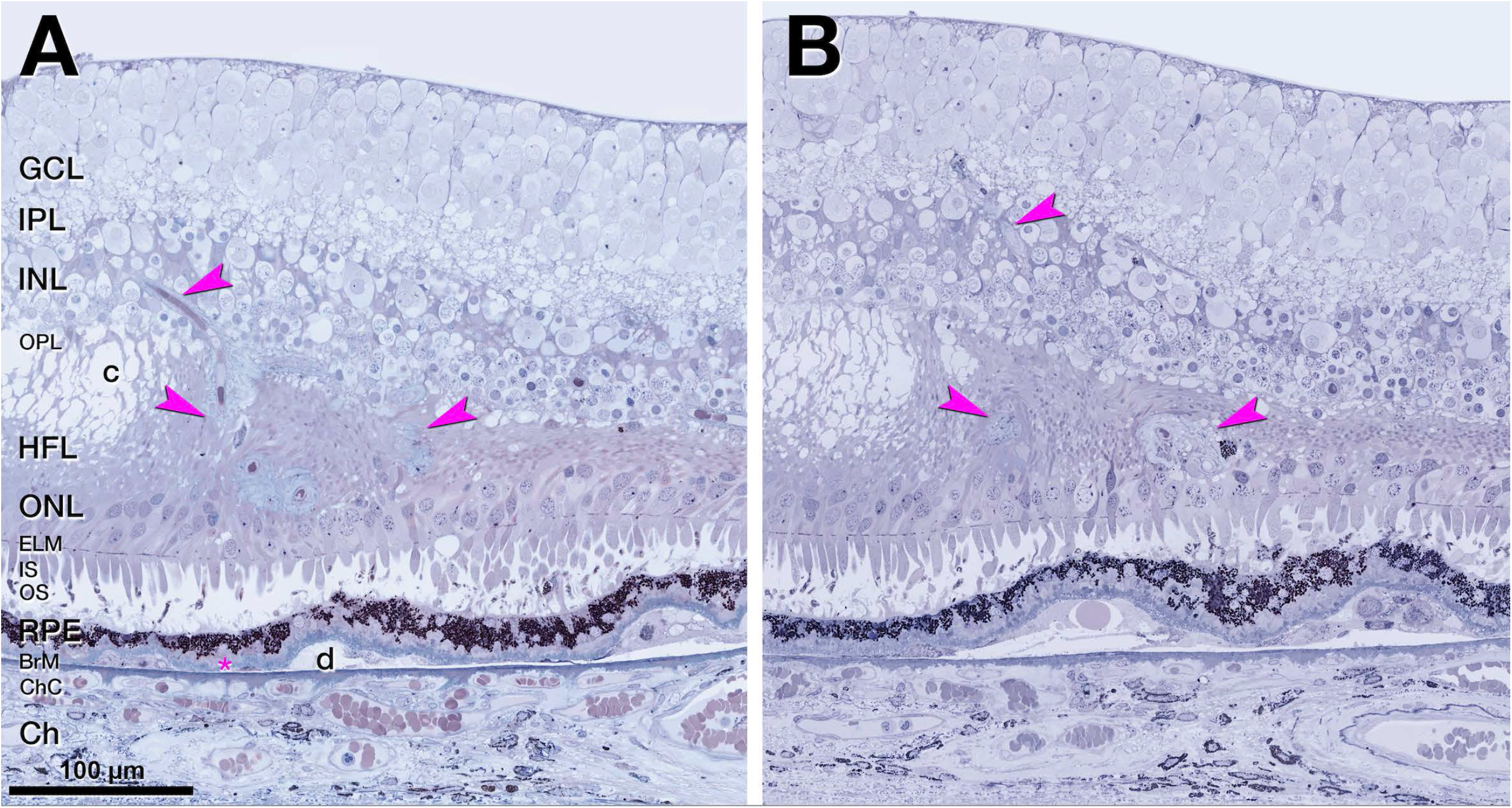
Vascular complex of deep retinal age-related microvascular anomaly (DRAMA), OD 3. **A, B**. A penetrating pair of vessels, (DRAMAs, fuchsia arrowheads) ensheathed by collagenous material dives from the inner nuclear layer (INL) into the Henle fiber layer (HFL)/outer nuclear layer (ONL). Next to the vessels is a degenerative cyst in the Henle fiber layer (HFL). There is no subsidence of external limiting membrane (ELM). The ONL is thinned. **B**. Retinal pigment epithelium (RPE) from the edge of a druse (d) migrates towards the lower edge of the vessel complex, right side.

### Vessel diameters of vascular lesions and comparison vessels

**Table S3** shows internal and external vessel diameters of type 3 MNV and DRAMA, as well as DCP vessels in the index case and donor eyes (8 intermediate AMD, 83.4 ± 11.6 years; 8 controls, 84.1 ± 6.7 years). Five of the 6 index case lesions are noticeably larger (2.5-3-fold) than the comparison vessels. External vessel diameters of type 3 MNV (OD 1, 15.02 µm ± 3.81 µm; OD 2, 21.35 µm ± 10.79 µm; OS 4 12.46 µm ± 0.95 µm) are larger than nearby DCP diameters (7.18 µm ± 1.11 µm). They are also larger than vessel diameters in intermediate AMD (6.79 µm ± 1.05 µm) and control (7.86 µm ± 1.47 µm, **Table S3A**) eyes. In DRAMA, external vessel diameters of OS 1 and OS 3 (13.39 µm ± 2.68 µm; 17.97 µm ± 1.08 µm) are considerably larger than the comparison vessels. A similar pattern is seen for internal vessel diameters (**Table S3B**).

An overview of unifying and distinguishing features of type 3 MNV and DRAMA, combining this and prior reports, is provided in **Table 4**.^3, 16, 24^

**Table 4.** Features of Type 3 MNV and DRAMA compared.

| **Feature** | **Type 3 MNV pyramidal** | **Type 3 MNV tangled** | **DRAMA** |
| --- | --- | --- | --- |
| Location, topographic | ETDRS inner | ETDRS inner | ETDRS inner |
| Location, layer | HFL/ONL | HFL/ONL | HFL/ONL |
| RPE | Absent, scattered, migrated | Absent, scattered, migrated | Migrated, intact |
| ELM | descent | descent | horizontal |
| Sub-RPE-BL space | End-stages of soft drusen* | End-stages of soft drusen* | Soft drusen, cells |
| Originating plexus | DCP | DCP | DCP |
| Shape of vascular complex | Compact, base-down pyramid | Vertical, horizontally spreading | Downward extending loop |
| Structure over time | dynamic | dynamic | stable<sup>24</sup> |
| Pericytes | Yes | Yes | Yes |
| Exudation | Yes | Yes | No |
| Anti-VEGF response | Yes | Yes | No |
| Collagenous sheath | Thick | Thin | None/thin if advanced |
| Endothelium | Non-fenestrated | Non-fenestrated | Non-fenestrated |

\*Calcific nodules, cells (subducted RPE, macrophages, giant cells, fibroblasts), Müller glia, avascular fibrosis Based on current and published data.<sup>3, 16, 24</sup>

## Discussion

In two anti-VEGF treated eyes of one patient with neovascular AMD, we compare longitudinal multimodal clinical imaging and histology of type 3 MNV and DRAMA (**Table 2, Table 4**). Although one patient, the clinical and imaging characteristics are typical of multifocal type 3 MNV, including bilaterality (55% of cases)^44^ and absence of type 1 MNV.^11-13, 44, 45^ Type 3 MNV is distinguished by intraretinal origin,^1, 2, 46^ frequent near-term bilateral involvement,^7-9^ and significant choroidal thinning with reduced perfusion.^47, 48^ Patients with type 3 MNV are older at initial diagnosis than patients with type 1 MNV.^49, 50^

In our case, treated type 3 MNV has two morphologic phenotypes, pyramidal (OD 2, OS 4) or tangled (OD 1). All three analyzed complexes originate at the DCP and extend posteriorly to approach persistent BLamD but do not enter the sub-RPE-BL space or cross BrM. These two phenotypes may correspond to those seen by Borelli et al in treatment-naïve type 3 MNV eyes with rotational three-dimensional OCTA. These authors describe 26 lesions as ‘filiform’ and 9 lesions as ‘saccular’, which appear similar in shape to pyramidal (filiform) and tangled (saccular).^14^ We also find more pyramidal lesions than tangled. It remains to be determined if saccular and filiform lesions differ in spatial distribution and time of onset. ^14^

Our previous description of pyramidal Type 3 MNV included endothelial cells in a thick collagenous matrix and dysmorphic RPE cells scattered along the neovascular stalk. ^16^ OD2 and OS4 in this case ^3^ add pericytes plus a nearly continuous covering by highly pigmented RPE (OS4), supporting the early involvement of migratory RPE.^2, 51^ The sheath distinguishes lesions from unaffected DCP vessels. The main differences between pyramidal and tangled lesions are the larger horizontal extent and thinner collagenous sheath of tangled.^3^ Endothelial cells of tangled vessels, and presumably also those in pyramidal, lack fenestrations like the source vessels in the DCP.

As before,^16^ we did not see vessels of choroidal origin in the sub-RPE-BL space, as reported for advanced type 3 MNV. ^14, 44, 52-54^ Nor did we see DCP-originating vessels penetrate through BLamD and enter the sub-RPE-BL space. This depth of penetration was suggested as necessary for exudation.^55^ This proposal was based on OCTA imaging without projection artifact removal to reduce spurious signal directly under type 3 MNV.^55^ Using volumetric artifact removal, we did not see OCTA flow signal in the sub-RPE-BL space (**Figure 8A3**). It is possible that anti-VEGF treatment rendered invisible downward projections from the DCP or upward projections from the choriocapillaris. We think this is unlikely, because neo-capillaries may shrink after treatment, but they do not disappear.^56, 57^ Further, our use of stepped sections may miss key details. Nevertheless, in all analyzed sections of this case, both BLamD and BrM were intact. Longitudinal imaging before exudation onset is needed to understand this phase of type 3 MNV progression.

The thick collagenous sheath of type 3 MNV lesions may impact clinical monitoring, as follows. All lesions responded to anti-VEGF with temporary resolution of exudation and persistence over several fluid absorption cycles. By OCTA, some early-stage lesions regress completely while treatment of later-stage lesions require continuing therapy.^2, 54^ Further, treated type 3 MNV lesions can reappear after becoming undetectable on OCTA. ^55, 58^ Lesions with thick sheaths remain detectable by structural OCT long after treatment.^57^ The collagenous neovascular stalk was populated by endothelial cells and pericytes that either remained, self-renewed, or migrated in during VEGF cycles. We can speculate that the sheath structurally stabilizes endothelial cells within it and that both VEGF and anti-VEGF agents impact lesions principally where the sheath is absent. If MNV persistence after treatment, as well as recurrence after apparent regression, is influenced by sheaths, then monitoring for disease activity by vascular characteristics in addition to fluid may be useful.

We showed that type 3 MNV can coexist with vascular formations that are candidate precursors, early stages, and masqueraders for type 3 MNV. All three DRAMA originated from the DCP and extended posteriorly into the HFL. None of the DRAMA lesions involved the superficial capillary plexus, and no significant intraretinal fluid was detectable on OCT over time. Like type 3 MNV (**Figure 10**), one DRAMA had non-fenestrated endothelial cells and pericytes (**Figure S14**). Unlike type 3 MNV and like native DCP, two DRAMA lacked a collagenous sheath. The one DRAMA with a thin sheath (OD 3) also extended furthest of the three into the ONL, perhaps indicating chronicity. Importantly, none of the DRAMA were accompanied by descent of the ELM, the border of atrophy in neurosensory retina, although the ELM can be perforated by inwardly migrating RPE as in OS 3. It is unlikely that DRAMA in our case represent exudative or non-exudative perifoveal vascular anomalous complexes (ePVAC/ nePEVAC),^31,35,36^ which typically appear above the DCP. Microvascular anomalies in eyes with neovascular and non-neovascular AMD include capillary dilations and telangiectasia that are associated with locally increased VEGF expression. ^17,59-61^ A recent study showed that 19/94 eyes with type 3 MNV exhibited an asymptomatic precursor stage on OCT.^44^ Because lack of exudation and non-progression to type 3 MNV in our case might result from VEGF suppression,^62^ longitudinal imaging is required to definitively place DRAMA in the progression sequence of type 3 MNV.

All analyzed vessels localized within 500-1500 µm of the foveal center, aligned with similar findings for solitary and multifocal lesions. ^11, 12^ A role for choroidal ischemia is hypothesized,^11, 63-65^ because the choroid is thinner in eyes with type 3 MNV than in eyes with types 1 and 2 MNV.^66, 67^ The radial symmetry of lesions around and close to the fovea further suggests an association with the distribution of photoreceptors and their support cells, which vary markedly in this eccentricity range. The ETDRS inner ring of type 3 MNV vulnerability is just peripheral to the foveal avascular zone^33^ on the inner slope of the crest of high rod density. Rod vision in AMD eyes is poorest in the same area. ^68-72^ Metabolic demand of foveal cones is high, and choriocapillaris OCTA signal decreases under the fovea throughout adulthood.^73, 74^ It is thus possible that the distribution of type 3 MNV is an additional effect of microvascular changes under the fovea that also contribute to high-risk drusen and reduced sustenance of nearby rods.

Study strengths include the availability of OCTA with eye-tracked OCT, volumetric projection artifact removal, rapid tissue preservation to largely maintain retinal attachment, registration of pre-mortem and post-mortem OCT volumes, and comprehensive histologic and microscopy techniques to reveal vessels and perivascular tissue elements. Limitations include the lack of longitudinal OCTA imaging ^9, 75^ and lack of color fundus photography to reveal discoloration patterns typical of type 3 MNV.^14^ Limitations to the laboratory study included use of stepped sections, lack of electron microscopy for all lesions, and lack of immunohistochemistry to support cell type identifications based on morphologic criteria. Finally, observations from one patient however detailed cannot elucidate the full range of biologic variability.

Nevertheless, our study helped define morphologies of type 3 MNV and proposed precursors that might guide future research, diagnosis, and disease monitoring. The presence and extent of a collagenous sheath distinguishes type 3 MNV from normal DCP vessels and may represent a stage in the evolution of DRAMA toward type 3 MNV. Our hypotheses can be tested in the larger samples available in clinic populations and clinical trial imaging datasets, ideally before onset of exudation.

## Data Availability

All data produced in the present study are available upon reasonable request to the authors

## Abbreviations

AMD: age-related macular degeneration
BLamD: basal laminar deposit
BrM: Bruch’s membrane
ChC: choriocapillaris
Ch: choroid
DCP: deep capillary plexus
DRAMA: deep retinal age-related microvascular anomalies
ELM: external limiting membrane
ETDRS: Early Treatment Diabetic Retinopathy Study
FA: fluorescein angiography
GCL: ganglion cell layer
HFL: Henle fiber layer
INL: inner nuclear layer
IPL: inner plexiform layer
IS: inner segment
MNV: macular neovascularization
nvAMD: neovascular age-related macular degeneration
OCT: optical coherence tomography
OCTA: optical coherence tomography angiography
ONL: outer nuclear layer
OPL: outer plexiform layer
OS: outer segment
RPE: retinal pigment epithelium.

## Contribution statement

All authors were involved in drafting the article or revising it critically for important intellectual content, and all authors approved the final version to be published. Dr. Curcio and Dr. Berlin had full access to all the data in the clinical picture and take responsibility for the integrity of the data and the accuracy of the data analysis. Study conception and design: AB, DC, LC, CB, RM, DF, KBF, CC. Acquisition of data: RM, CB, JM, LC, AB, DC, KBF, CC. Analysis and interpretation of data: AB, DC, LC, DF, KBF, CC. Writing of manuscript: AB, DC, CB, LC, RM, DF, KBF, CC.

## Meeting presentation

This work was submitted as part of an abstract to the annual meeting of The Association for Research in Vision & Ophthalmology (ARVO) May 2021 and May 2022.

## Figures

**Figure S2.**
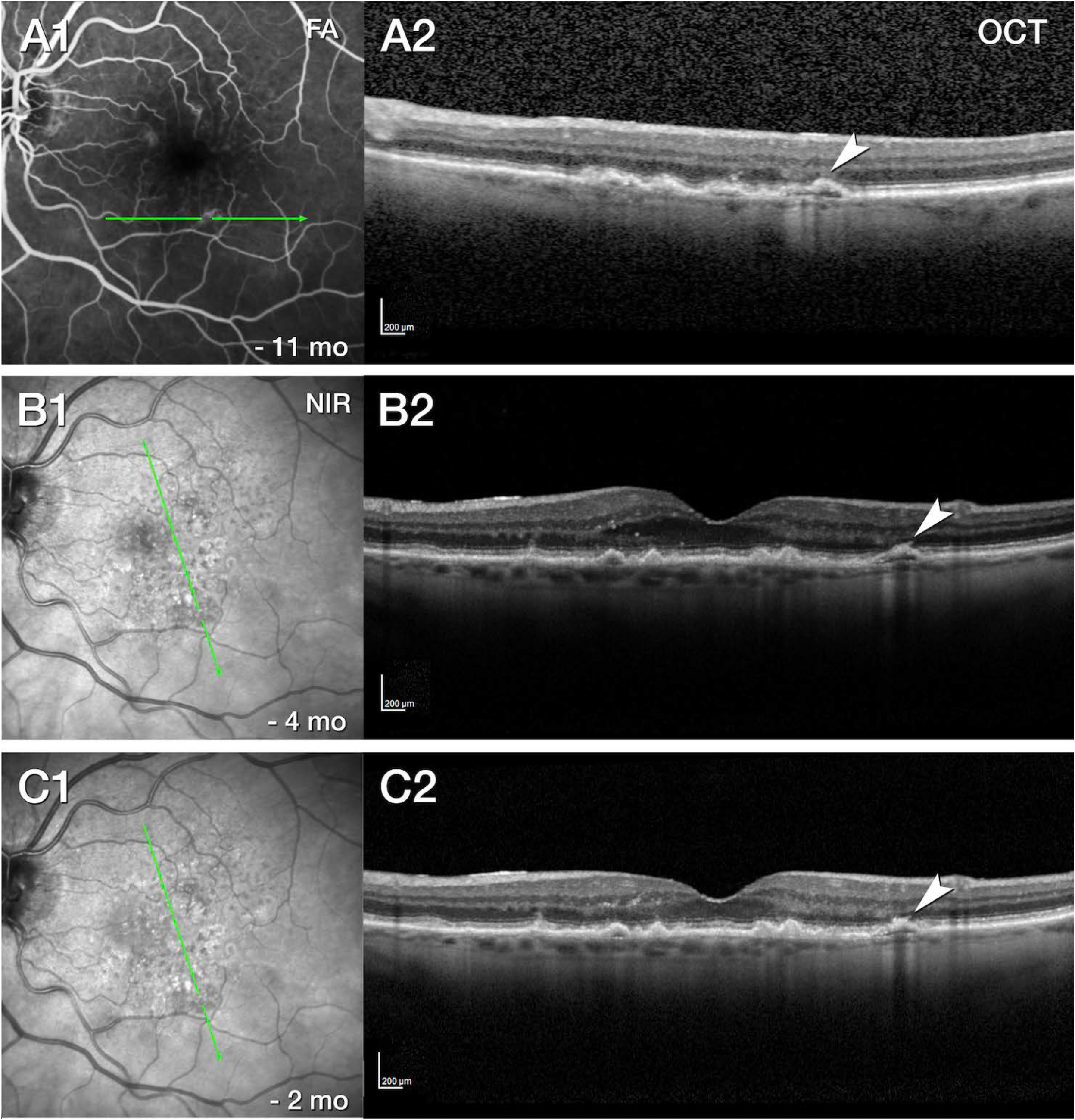
Hyperfluorescence without histologic correlate, OS 2. **A**. Fluorescein angiography (FA, **A1**) venous phase, 11 months before death, shows multiple instances of hyperfluorescence. An optical coherence tomography (OCT) B-scan (green line) displays drusen, a double-layer sign (white arrowhead), choroidal hypertransmission, and hyperreflective foci (**A2**). **B, C**. At 4 and 2 months before death, with 5 (**B**) and 6 (**C**) total injections, respectively, the lesion is stable, without intraretinal cysts.

**Figure S3.**
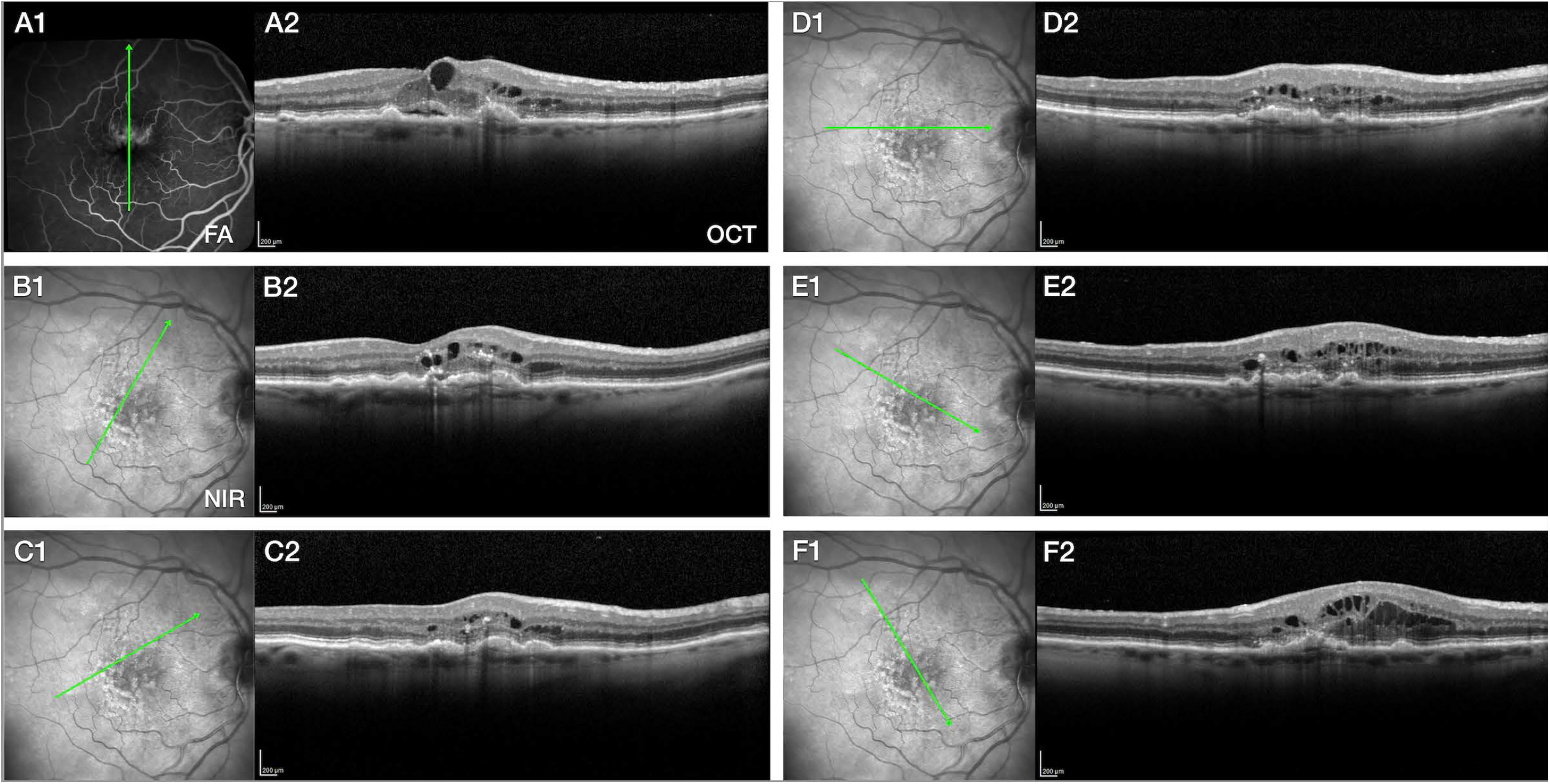
Initial presentation of right eye, 5 years before death. **A-E**. Fluorescein angiography (FA), near infrared reflectance images (NIR), and radial optical coherence tomography (OCT) B-scans show exudative age-related macular degeneration in the right eye at initial presentation. Based on these findings anti-VEGF therapy is initiated. **A**. FA recirculation phase shows marked leakage. Corresponding OCT shows intraretinal and subretinal hyporeflective cysts and spaces representing exudation, located above drusenoid pigment epithelium detachments in the fovea and parafovea. **B**. NIR and OCT show hyporeflective subretinal drusenoid deposits and hyperreflective soft drusen across the macula. Hyperreflective foci are present in inner and outer nuclear layer.

**Figure S4.**
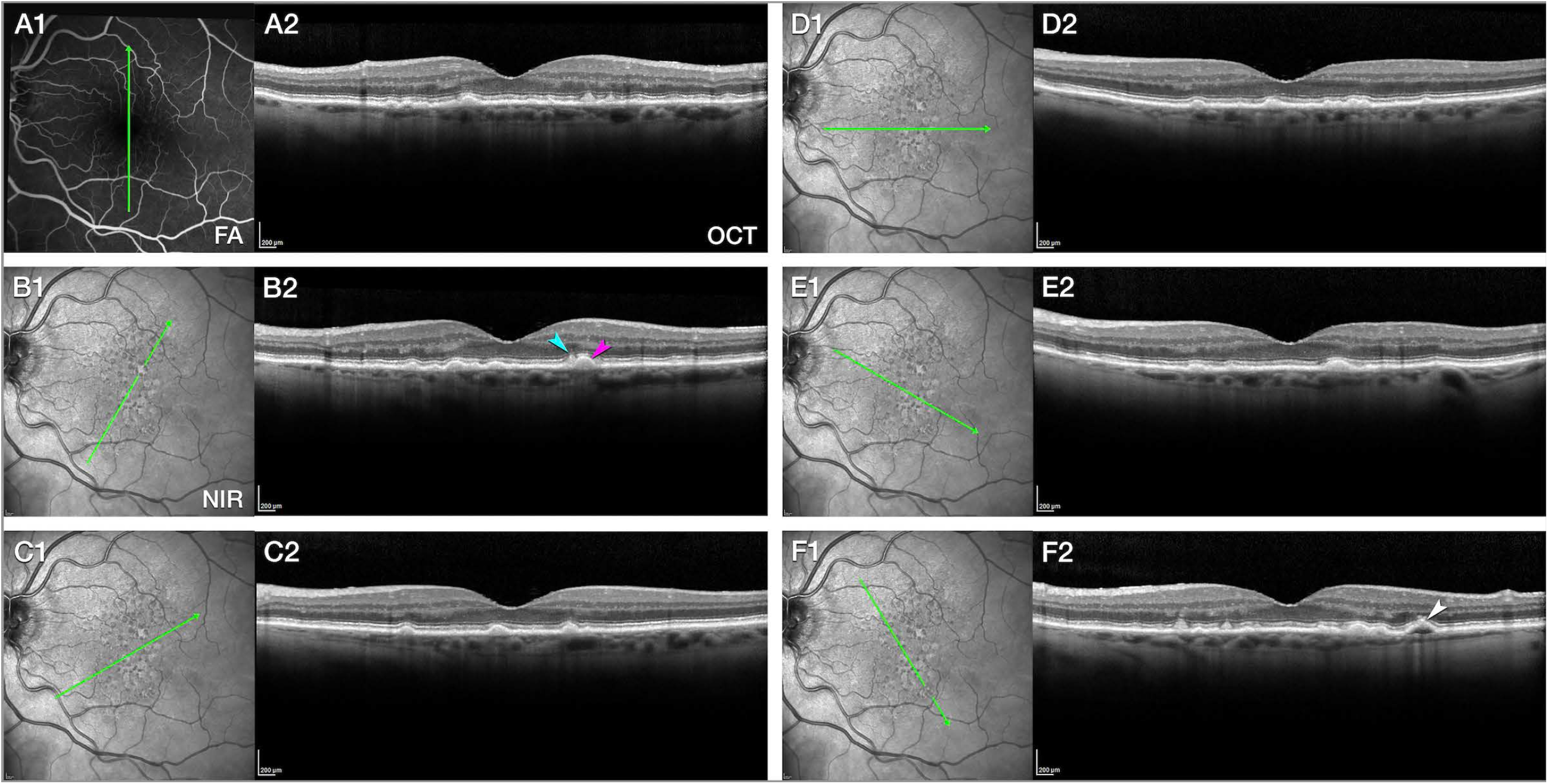
Initial presentation of left eye, 5 years before death. **A-E**. Near infrared reflectance images (NIR) and radial optical coherence tomography (OCT) B-scans show non-exudative age-related macular degeneration in the left eye. Hyporeflective subretinal drusenoid deposits and hyperreflective soft drusen (**B** teal and fuchsia arrowheads respectively) appear across the macula. **B**. On the recirculation phase of fluorescein angiography (FA), no intraretinal or subretinal fluid or leakage is present, ensuring absence of active MNV exudation. **F**. A shallow RPE elevation with choroidal hypertransmission (white arrowhead) and hyperreflective focus was stable on longitudinal follow-up by OCT (**Figure S4**). No vessel was found in histology. This lesion presumably corresponds to a calcified druse.^39^

**Figure S10.**
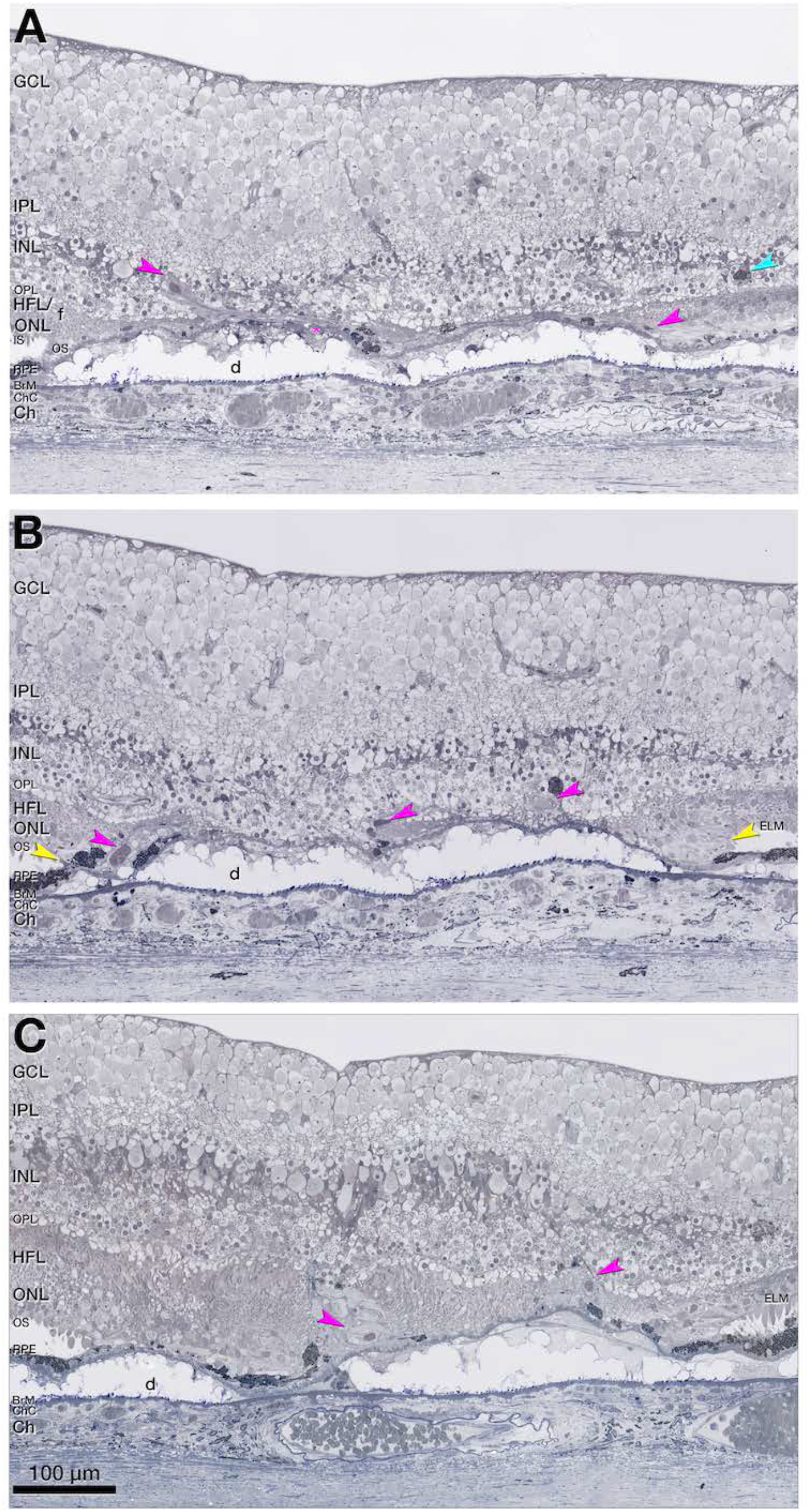
Tangled vascular complex in type 3 MNV, OD 1. **A-C**. Tangled vascular complex (fuchsia arrowheads indicate lumen) spans 249 µm horizontally towards the superior perifovea (**A**, 759 µm from fovea; **B**, 719 µm from fovea; **C**, 552 µm from fovea). The complex is partly ensheathed by collagenous material and is flanked by retinal pigment epithelium (RPE) cells. Two cells rest entirely within the outer plexiform layer (OPL)/ inner nuclear layer (INL; light blue arrowhead in **A**). The INL/OPL subsides, and the vascular complex extends from the INL/OPL border through the Henle fiber layer (HFL)/ outer nuclear layer (ONL). The complex adheres to basal laminar deposits (BLamD) draping a calcified druse (d). Bruch’s membrane (BrM) appears intact without evidence of a choroidal contribution. The external limiting membrane (ELM) descends at both edges of the calcified druse (yellow arrowheads in **B**). Vessel walls do not exhibit obvious arterial or venous features. Vessel diameter within the INL was larger than 15 µm, suggesting drainage venules.

**Figure S14.**
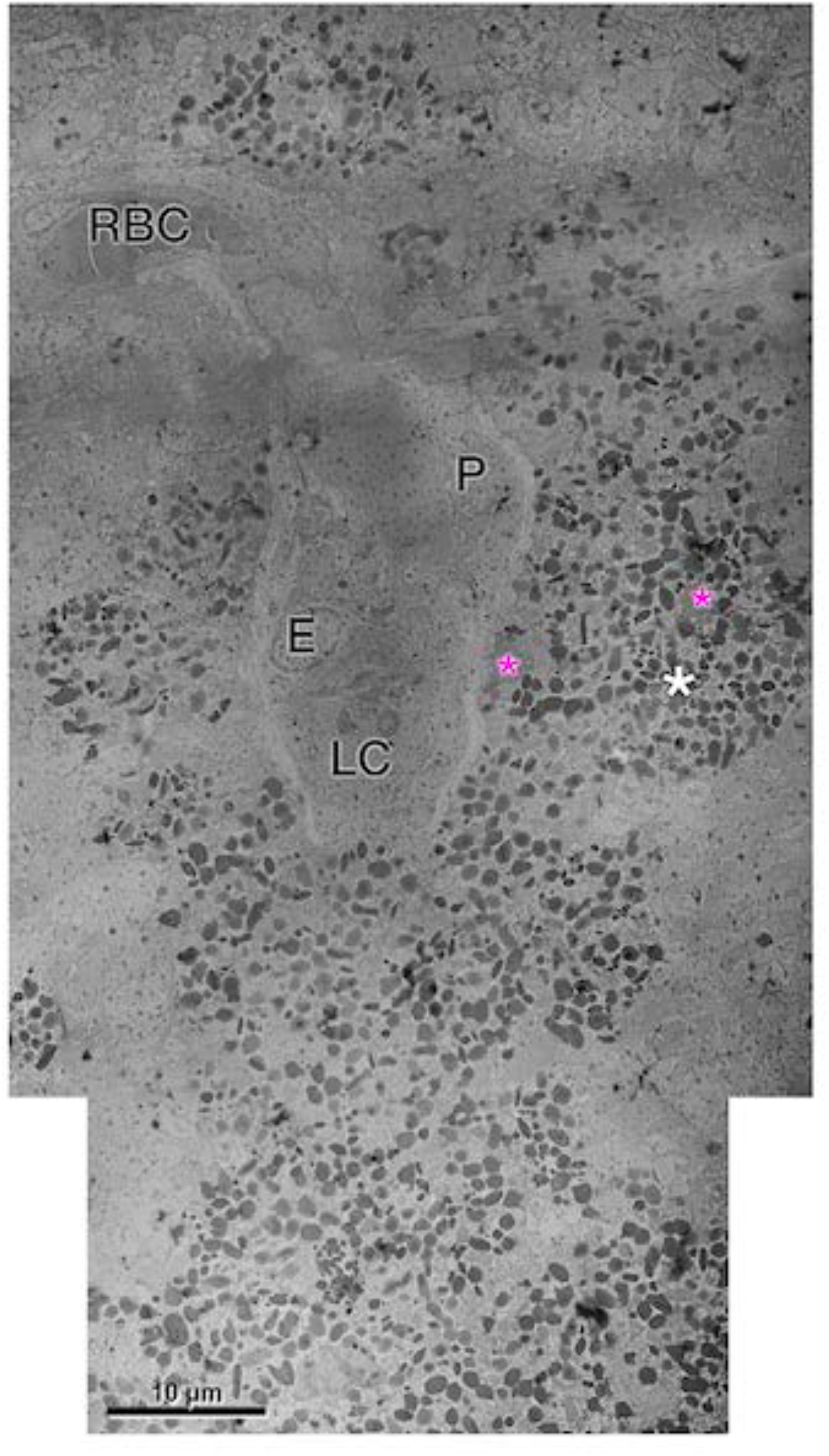
Transmission electron microscopy of DRAMA with RPE complex, OS 3. The Vessel is ensheathed by an endothelial (E) cell and a pericyte (P), with little collagen. The Lumen contains a leukocyte (LC) and a red blood cell (RBC). The surrounding tower of retinal pigment epithelium (RPE) is multicellular (white asterisk), and multinucleated (fuchsia asterisks). RPE organelle packing density and electron-density is similar to in-layer RPE cells (not shown).

**Video S1**.

**Volume rendering of structural OCT (gray) and OCTA (yellow) of tangled type 3 MNV, OD 1**.

The cube is rotated 180° to highlight vascular findings in the superior macula region. Coronal view demonstrates blood flow immediately below the deep capillary plexus level in two separate areas. Superposition with structural OCT (gray channel) demonstrates neovascular blood flow within an hyperreflective structure at the outer nuclear layer and above Bruch’s membrane (BrM). The superficial arteries (red) and veins (blue) in the vicinity of the lesion are outlined after fluorescein angiography analysis. Three-dimensional analysis of neovascular blood flow depicts an anastomosis right above BrM (green section) Spinning around the neovascular blood-flow highlights separate inflow and outflow.

---

**Table S1.** Content of imaging data pieces.

| figure number as appearing | Laterality and lesion number | Lesion type | Content | Title |
| --- | --- | --- | --- | --- |
| **1** | NA | NA | multimodal imaging | Multimodal retinal imaging of both eyes, 11 months before death |
| **S2** | OS2 | type 3 MNV | Clinicopathologic correlation, panoramic histology | Hyperfluorescence without histologic correlate, OS2. |
| **S3** | NA | NA | clinical imaging baseline | Initial presentation of right eye, 5 years before death |
| **S4** | NA | NA | clinical imaging baseline | Initial presentation of left eye, 5 years before death |
| **5** | OS4 | type 3 MNV | Clinicopathologic correlation, panoramic histology | Multimodal imaging, clinical course, and histology of pyramidal type 3 MNV, OS4. |
| **6** | OS4 | type 3 MNV | Magnified histology | <u>Pyramidal vascular complex in type 3 MNV, OS4.</u> |
| **7** | OS4 | type 3 MNV | Transmission electron microscopy | Transmission electron microscopy of pyramidal type 3 MNV, OS4 |
| **8** | OD1 | type 3 MNV | Clinicopathologic correlation, panoramic histology | Multimodal imaging, clinical course, and histology of tangled type 3 MNV, OD1. |
| **S9** | OD1 | type 3 MNV | 3D Volume rendering | Volume rendering of structural optical coherence tomography (OCT, gray) and OCT angiography (OCTA, yellow) of tangled type 3 MNV, OD1. |
| **S10** | OD1 | type 3 MNV | Magnified histology | Tangled vascular complex in type 3 MNV, OD1. |
| **11** | OS1 | DRAMA/<br>type 3 MNV<br>precursor | Clinicopathologic<br>correlation,<br>panoramic<br>histology | Deep retinal age-related microvascular anomaly (DRAMA),<br>OS1. |
| **12** | OS3 | DRAMA/<br>type 3 MNV<br>precursor | Clinicopathologic<br>correlation,<br>panoramic<br>histology | Multimodal imaging and histology of deep retinal age-related<br>microvascular anomaly (DRAMA) with intraretinal RPE<br>complex, OS3. |
| **13** | OS3 | DRAMA/<br>type 3 MNV<br>precursor | Magnified<br>histology &<br>Transmission<br>electron<br>microscopy | RPE complex associated with deep retinal age-related<br>microvascular anomaly (DRAMA), OS3. |
| **S14** | OS3 | DRAMA/<br>type 3 MNV<br>precursor | Transmission<br>electron<br>microscopy | Transmission electron microscopy of DRAMA with RPE<br>complex, OS3. |
| **15** | OD3 | DRAMA/<br>type 3 MNV<br>precursor | Clinicopathologic<br>correlation,<br>panoramic<br>histology | Multimodal imaging of deep retinal age-related microvascular<br>anomalies (DRAMAs), OD3. |
| **16** | OD3 | DRAMA/<br>type 3 MNV<br>precursor | Magnified<br>histology | Vascular complex of deep retinal age-related microvascular<br>anomaly (DRAMA), OD3. |

T3MNV, type 3 macular neovascularization; DRAMA, deep retinal age-related microvascular anomaly; OD, right eye; OS, left eye.

**Table S3.** Diameters of vessels in histology.

**A. External vessel diameter (μm)**
|  | **T3 MNV\*** |  |  | **DRAMA\*** |  |  | **T3 DCP†** | **Control†** | **AMD†** |
| --- | --- | --- | --- | --- | --- | --- | --- | --- | --- |
|  | OD 1 | OD 2 | OS 4 | OS 1 | OS 3 | OD 3 |  |  |  |
| Mean | 15.02 | 21.35 | 12.46 | 13.39 | 17.97 | 8.62 | 7.18 | 7.86 | 6.79 |
| SD | 3.81 | 10.79 | 0.95 | 2.68 | 1.08 | 1.05 | 1.11 | 1.47 | 1.05 |

**B. Internal vessel diameter (μm)**
|  | **T3 MNV\*** |  |  | **DRAMA\*** |  |  | **T3 DCP†** | **Control†** | **AMD††** |
| --- | --- | --- | --- | --- | --- | --- | --- | --- | --- |
|  | OD 1 | OD 2 | OS 4 | OS 1 | OS 3 | OD 3 |  |  |  |
| Mean | 9.34 | 14.86 | 9.87 | 9.40 | 8.28 | 5.17 | 3.69 | 4.44 | 3.75 |
| SD | 1.85 | 6.19 | 0.76 | 2.00 | 0.31 | 0.34 | 0.92 | 0.98 | 0.92 |

\* 3 to 6 measurements for each; T3 MNV, type 3 macular neovascularization; DRAMA, deep retinal age-related microvascular anomaly.
† 8 female control eyes (84.1 ± 6.7 years; 1 male and 7 female), intermediate AMD eyes (83.4 ± 11.6 years). Histology from Project MACULA https://projectmacula.org.
Number of measurements, Type 3 DCP (n=123); Controls (n=88); AMD (n=107).
OS2 no lesion identified in histology.

